# Sex adaptive deep recurrent neural networks for Parkinson’s disease detection using 5-second vertical ground reaction force signals

**DOI:** 10.1101/2025.11.01.25339121

**Authors:** Gurkirtan Singh, Anilendu Pramanik

## Abstract

**Background:** This study introduces an innovative sex-stratified methodology for the identification of Parkinson’s disease (PD) using vertical ground reaction force (VGRF) data obtained from foot sensors during ambulation. We devised and evaluated four distinct recurrent neural network designs.

**Objectives:** We identified a substantial deficiency in existing diagnostic methodologies that infrequently utilize sex-stratified techniques by creating distinct models for male and female individuals over the age of 50 years and architectural benchmarking for variants of recurrent neural networks in processing vertical ground reaction force signals.

**Methodology:** We trained Long Short-Term Memory (LSTM), bidirectional LSTM (bi-LSTM), Gated Recurrent Unit (GRU), and bidirectional GRU (bi-GRU) on brief 5 s intervals of vertical ground reaction force (VGRF) data.

**Results:** Our results indicate that the GRU models, which use fewer resources, perform very well, achieving high accuracy (98.5% for males and 99.46% for females), recall (98.91% for males and 100% for females), and F1 scores (98.78% for males and 99.48% for females), compared to the more complex LSTM models. The bidirectional models demonstrated similar performance but necessitated increased processing resources.

**Conclusion:** The efficacy of these sex-specific models with abbreviated time frames underscores the possibility of more tailored, efficient, and accessible Parkinson’s disease screening instruments that could facilitate earlier intervention and treatment. Our discovery signifies a significant step in streamlining the diagnostic procedure for Parkinson’s disease patients non-invasively while also establishing a basis for the future development of real-time diagnostic in clinical devices.

## Introduction

Parkinson’s disease (PD) is a progressive neurological condition affecting approximately 10 million people worldwide. It is characterized by motor symptoms including bradykinesia, rigidity, and gait abnormalities, as noted by the Parkinson’s Foundation(1). The degeneration of dopaminergic neurons in the substantia nigra is a diagnostic indicator of Parkinson’s (Poewe et al.(2)). Research indicates that men experience Parkinson’s disease 1.5 times more frequently than women (Navita et al.(3)). Early identification is difficult because initial symptoms, especially gait patterns, may resemble age-related changes (Yin et al.(4), Veeraragavan et al.(5)). Vertical ground reaction force (VGRF) signals, obtained via wearable sensors during ambulation, have become a pivotal biomarker for Parkinson’s disease detection, providing non-invasive measurement of gait dynamics, including stride variability, weight distribution asymmetry, and temporal force patterns (Veeraragavan et al.(5), Khoury et al.(6)).

Conventional Parkinson’s diagnosis techniques depend significantly on subjective clinical evaluations, which are prone to inter-rater variability and exhibit poor sensitivity to early-stage symptoms (Yin et al.(4), Veeraragavan et al.(5)). Machine learning methodologies utilizing VGRF data mitigate these constraints by extracting objective spatiotemporal features, such as heel-strike impulse asymmetry and mid-stance force modulation, which are substantially correlated with dopaminergic degeneration (Veeraragavan et al.(5), Khoury et al.(6)). Recent research indicates classification accuracies of over 97% when employing artificial neural networks on VGRF datasets, underscoring its diagnostic potential (Veeraragavan et al.(5)).

Recurrent Neural Networks (RNNs) are excellent for analyzing VGRF time-series data because of their ability to represent temporal correlations in gait cycles. Long Short-Term Memory (LSTM) and Gated Recurrent Unit (GRU) architectures, along with their bidirectional variants (bi-LSTM/bi-GRU), excel at capturing forward and backward contextual relationships in sequential force patterns (Mir et al.(7), Skaramagkas et al.(8)). For instance, a study conducted by Skaramagkas et al.(8) showed bidirectional LSTM models with attention mechanisms achieving 98% accuracy in detecting subtle PD-related tremors from sensor data, outperforming conventional ML methods.

Sex-specific modeling is essential in PD identification, as it might uncover distinct gait adaptations between males and females. Contemporary techniques for PD detection rarely employ sex-stratified methodologies, which may diminish the sensitivity in diverse populations.

This research identifies three critical deficiencies:

Sex-adaptive modeling: building separate LSTM, bi-LSTM, GRU, and bi-GRU architectures for male and female cohorts to account for biomechanical differences in VGRF patterns.

Modeling for a shorter time window of 5 s to capture more meaningful temporal characteristics may further improve the accuracy of sex-specific modeling.

Architecture benchmarking: systematic comparison of RNN variant performances in capturing PD-specific temporal gait features.

By evaluating these architectures on sex-stratified VGRF data, this study aimed to optimize model selection for real-world PD screening applications, enabling earlier intervention and personalized therapeutic strategies.

### Literature Review

Researchers have widely employed ML and deep learning (DL) methodologies for the identification of Parkinson’s disease (PD), utilizing multiple qualitative and quantitative characteristics, resulting in numerous publications. Accelerated technical progress in sensor-based devices within the healthcare sector has enabled more precise assessment of diverse motor and non-motor disorders. We identified foot sensors that measure VGRF signals as the most resource-efficient sensors for analyzing gait patterns. We thoroughly reviewed studies that used machine learning and deep learning techniques on VGRF sensor data and other sensors to better understand the information related to our goals.

Khera et al.(9) developed a model for the classification and severity assessment of Parkinson’s specific to age and sex. They utilized multiple ML approaches, including k-nearest neighbors (k-NN), support vector machines (SVM), decision trees (DT), and random forests, and employed a recursive feature selection method to develop and execute the model on the gait dataset using a 10-fold cross-validation strategy. The SVM emerged as the most effective classifier, achieving an accuracy of 98.5% in the classification of PD, surpassing another generalized model, but demonstrating a lower accuracy of 94.02% in the severity evaluation of PD. Time-domain characteristics were recovered from the sensor readings by gait cycle segmentation, and the VGRF data were partitioned into individual strides for feature extraction. This approach complicates the development of real-time PD diagnosis using VGRF sensors because a robust data preprocessing pipeline must be established prior to the extraction of all time-domain variables, rendering their model challenging to implement in real-time clinical settings. Recurrent neural networks obviate the necessity for distinct feature extraction and frequently surpass standard ML models, such as SVM, in the acquisition of temporal information.

In a recent study, Navita et al.(3) developed a complex ML model using a two-step classification approach. Step 1 had a hyper-tuned random forest tree (RFT) to categorize participants into PD and non-PD categories, and the following, step 2, a hyper-tuned ensemble regressor (ER) model to predict illness severity utilizing the VGRF dataset from Physionet(10). They employed the synthetic minority over-sampling technique (SMOTE) to generate synthetic data to balance the minority class samples. They utilized a recursive feature elimination technique to identify the ideal feature set for enhancing model performance(3). Using the RFT model, they achieved impressive results in differentiating Parkinson’s disease (PD) patients from non-PD individuals, reporting an accuracy of 97.5% ± 2.1%, a sensitivity of 97% ± 2.5%, and an average specificity of 95% ± 2.2%2. In the second phase, the model was employed to evaluate disease severity, yielding an average accuracy of 96.4% ± 2.3% with loss reported as 0.065 ± 0.024 (mean absolute error) and 0.080 ± 0.06 (root mean square error)(3). SMOTE employs the K-NN algorithm with Euclidean distance to produce synthetic samples from the minority class training samples; however, it neglects the time-series characteristics of VGRF signals(11) and may struggle to distinguish between stance and swing phases, which exhibit extreme VGRF values. Consequently, the generated samples may not accurately represent the readings obtained from both cohorts. However, if they had incorporated sex stratification into their model, it might have yielded superior outcomes.

Mir et al. (12) proposed an innovative LSTM methodology for detecting freezing of gait, utilizing the VGRF dataset from Physionet(10) to develop a classification model. The model achieved an accuracy of 97.71%, a sensitivity of 99%, a precision of 98%, and a specificity of 96%. They employed the Rectified Linear Unit (ReLU) activation function rather than adhering to the standard LSTM architecture, which is designed to address vanishing and exploding gradient issues. Furthermore, employing ReLU as the activation function renders model training computationally expensive. The parameters of the participants’ age were not taken into account, which included three younger cohorts aged 36, 37, and 45 years, while the remaining data comprised elderly subjects aged 50 years or older. They did not explore alternative types of recurrent neural networks, such as Gated Recurrent Units, which provide performance similar to that of LSTMs and, in certain instances, superior performance while being less computationally intensive.

Khoury et al.(6) suggested an ML-based method to differentiate between patients with PD and healthy individuals using VGRF sensor data obtained from Physionet(10). They conducted experiments with multiple supervised ML models, including Naive Bayes (NB), K-NN, DT, SVM, RFT, and Gaussian Mixture Models (GMM), as well as unsupervised ML categorization using K-means. The performance criteria employed for the assessment of generated models included accuracy, recall, F1 score, and precision, utilizing the Leave-One-Out Cross-Validation (LOOCV) method. Their suggested methodology was helpful in distinguishing between PD and HC, achieving a suboptimal accuracy rate of 90% utilizing K-NN. The accuracy might have been enhanced by employing deep learning methodologies such as recurrent neural networks and their derivatives.

Setiawan et al.(13) suggested a paradigm for the detection and assessment of the severity of PD. The model was developed by emphasizing time-frequency-domain features, which were altered through Continuous Wavelet Transformation (CWT) to produce a spectrogram for enhanced visualization. Principal Component Analysis (PCA) was employed to augment model accuracy by identifying significant components from the extracted feature set. The VGRF dataset was utilized to construct DL models, supplemented by 10-fold cross-validation to enhance generalizability. Separate models were constructed for each contributing dataset within the VGRF dataset, and the datasets were also amalgamated to develop more generalized models. The datasets underwent pre-processing to incorporate signal windows of 10s, 15s, and 30s duration. The classification was conducted in two manners: binary classification distinguishing between PD and healthy subjects, and multiclass classification based on Hoehn & Yahr severity ratings, categorizing into PD Stage 2, PD Stage 2.5, PD Stage 3, and healthy subjects, resulting in four distinct groups. The optimal outcome for the classification of the merged dataset was attained with the ResNet Convolutional Neural Network, which achieved a modest accuracy of 94.58%. They did not use a hybrid model, which may have enhanced their prediction accuracy scores. They did not test a reduced window size of 5 s and stratified the VGRF dataset by sex, which may have enhanced their results. The proposed model incorporates additional complications in data transformations, which complicates its usability and hinders its scalability in real-time diagnosis.

Joshi et al.(14) proposed an “automatic non-invasive method for Parkinson’s disease classification” that was able to differentiate between Parkinson’s gait and healthy subject gait by blending wavelet analysis with an SVM model. A low accuracy score of 90.32 % was obtained by considering only one gait parameter while considering specific gait variables, whereas the db2 wavelet underperformed the Haar wavelet decomposition(14). The dataset had only 31 subjects, including 16 healthy and 15 PD subjects, rendering the model’s performance inappropriate for a larger population.

### Methodology

The suggested technique includes dataset description, exploratory data analysis, data preparation, proposed models, model building, and evaluation metrics. We executed all phases of the suggested methodology utilizing the "Google Colab" Jupyter Notebook with Python version 3.11.11. The recommended methodology for the model-building process is illustrated in **Error! Reference source not found.**.

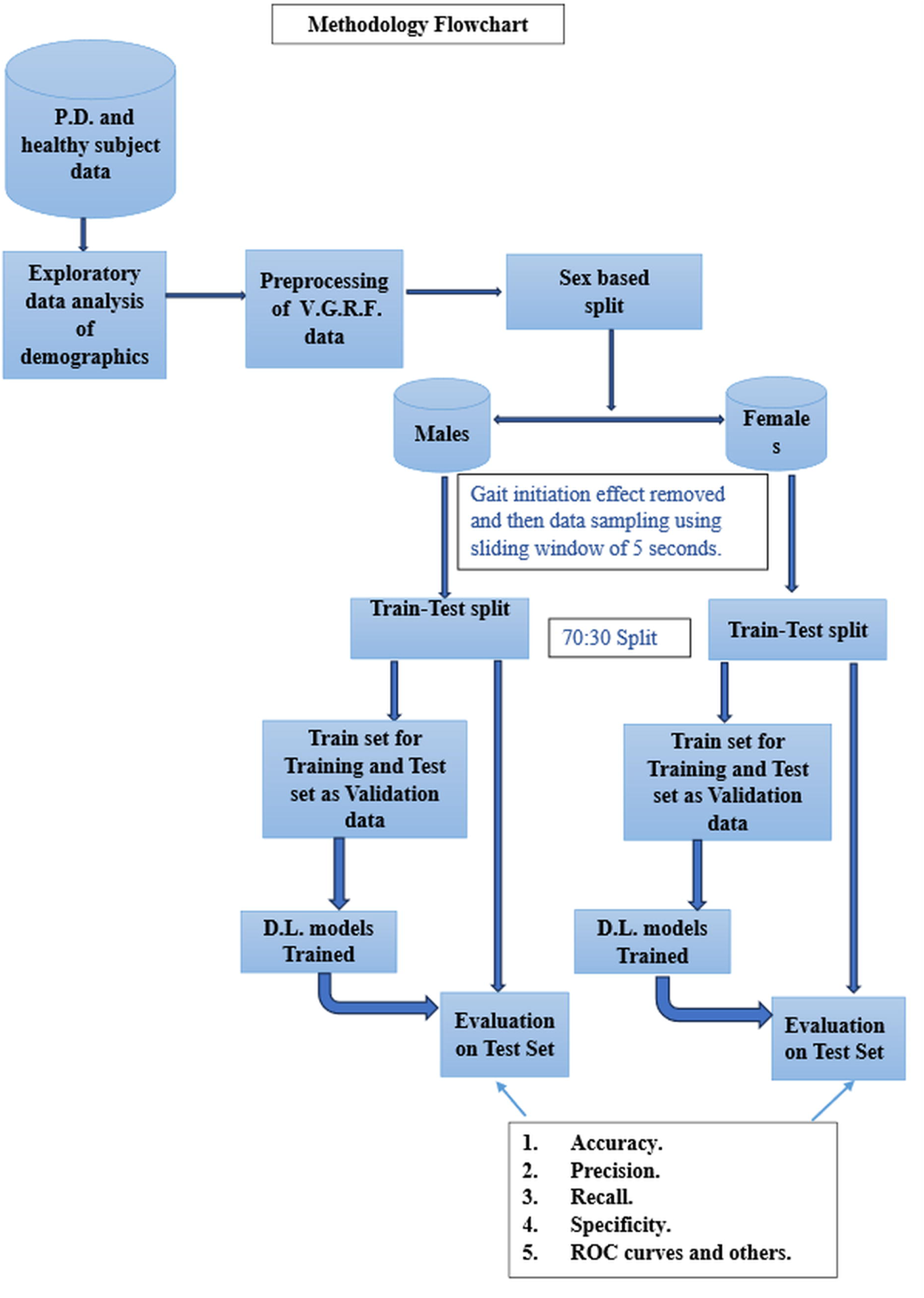

### Dataset Description

The dataset utilized for the implementation of the proposed models was obtained from Physionet(10).A secondary dataset was obtained for model development, which could be augmented using newly collected smart sensor data. The dataset was compiled from gait recordings obtained from the Movement Disorders Unit, Laboratory for Gait and Neurodynamics, Tel-Aviv Sourasky Medical Center. It was named in recognition of the researchers who contributed to its collection: Frenkel-Toledo et al.(15) (labeled as ‘Si’), Hausdorff et al.(16) (labeled as ‘Ju’), and Yogev et al.(17) (labeled as ‘Ga’). This dataset consists of gait signals collected from 166 individuals, including 93 diagnosed with PD and 73 categorized as healthy controls. We obtained a dataset containing demographic information for exploratory data analysis and cleansing. Figure 2, adapted from the study by Alam et al.(18), shows the layout of the sensor placements when a subject is standing in a relaxed position with their feet parallel. The approximate (X, Y) coordinates are provided relative to a coordinate system centered between the legs, with the person facing in the direction of the positive Y-axis.

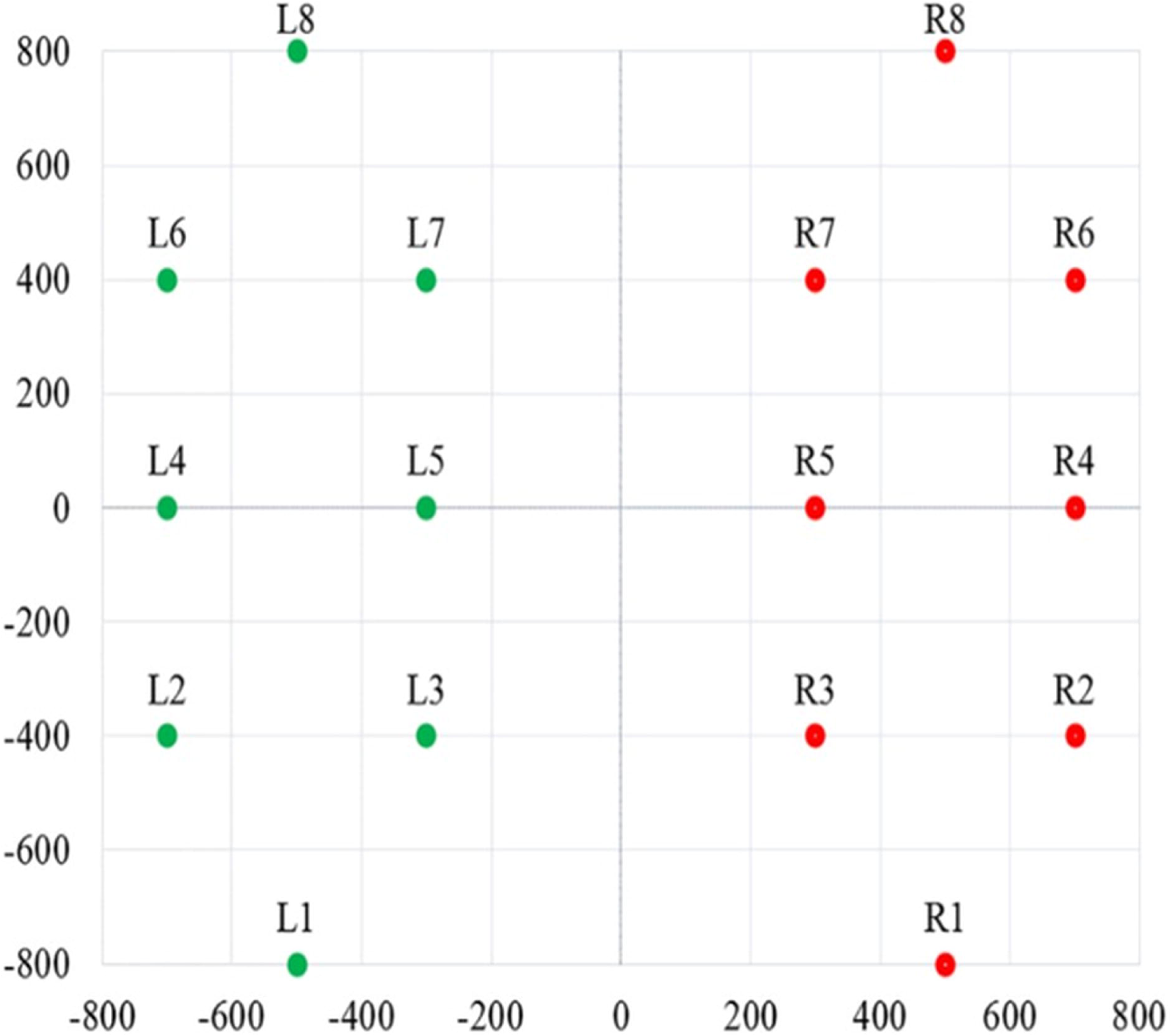

### Exploratory Data Analysis

The demographic data indicate the number of subjects from each study categorized by sex, as illustrated in Figure 3. The Hoehn and Yahr score for PD subjects was “2” for 55 subjects, “2.5” for 10 subjects, and 3 for 18 subjects. This range signifies progression from early-stage Parkinson’s (stages 1 and 2) to middle-stage disease, characterized by a more pronounced impact on daily functioning due to the onset of bilateral motor involvement, although independence is still preserved (Hoehn et al.(19)). The age distribution of the subjects, as illustrated by the boxplot in Figure 4, indicated significant outlier values. The dataset comprised sensor readings from both subject groups organized as separate text files with the attributes outlined in Table 1. Additionally, looking at the readings from each study showed significant differences in how long data was collected, as shown in Figure 5. Analysis of the "Total Force Under Left Foot" and "Total Force Under Right Foot" from the 10-second to 15-second interval of a randomly selected subject from each study group revealed significant fluctuations in peak force achievements over time, as also emphasized by the research conducted by Navita et al.(3) Figure 6 illustrates the related graphs.

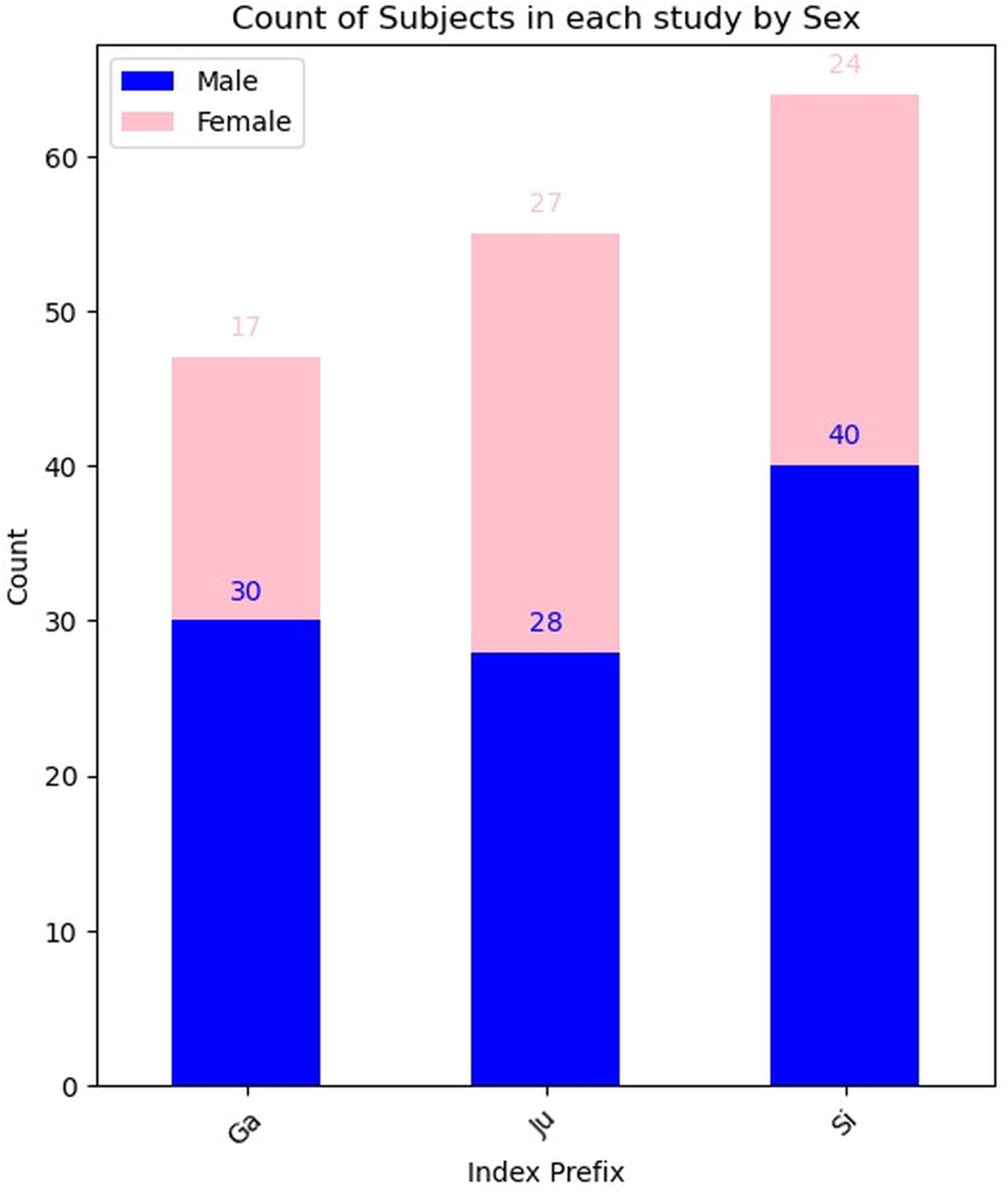

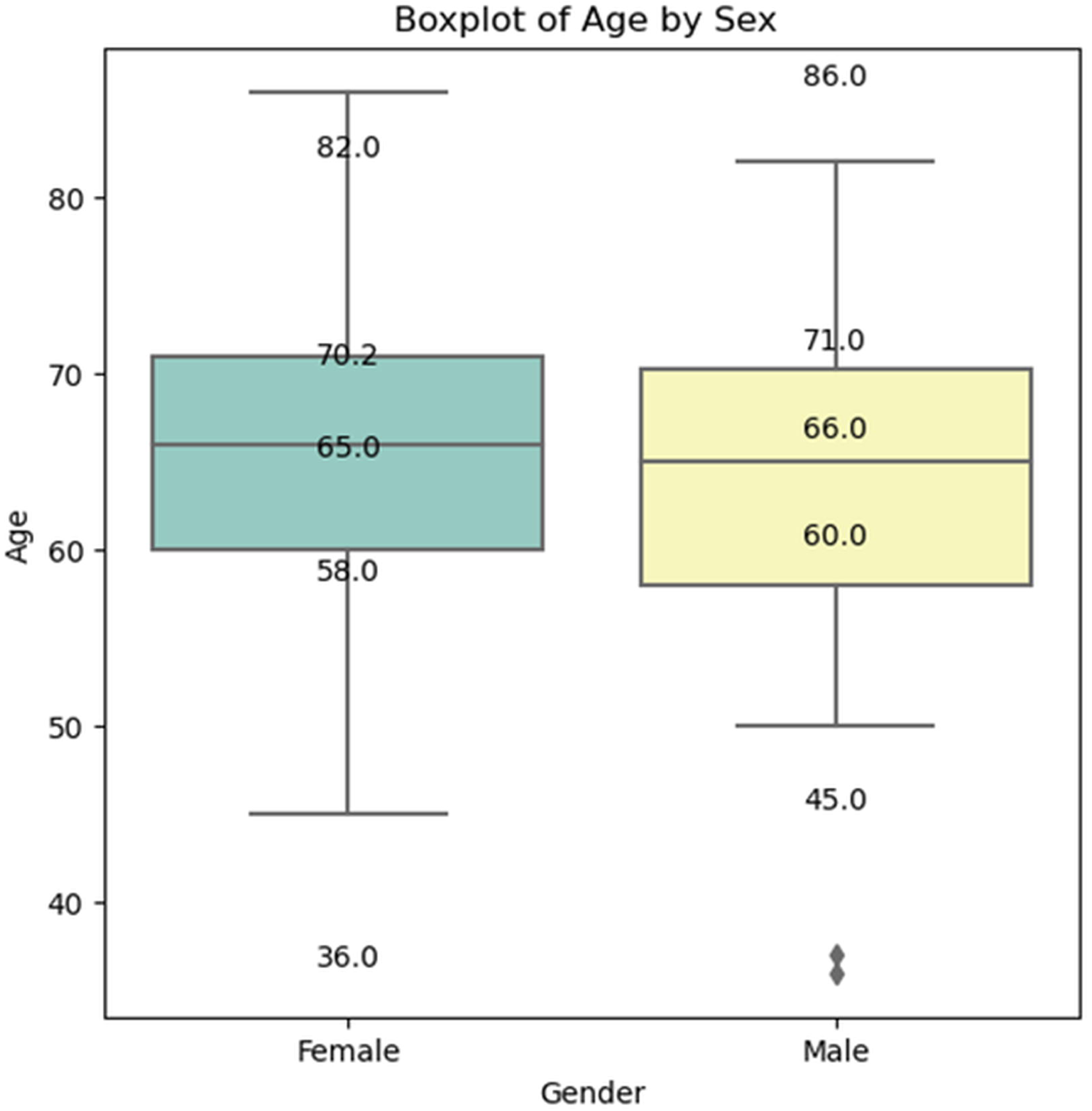

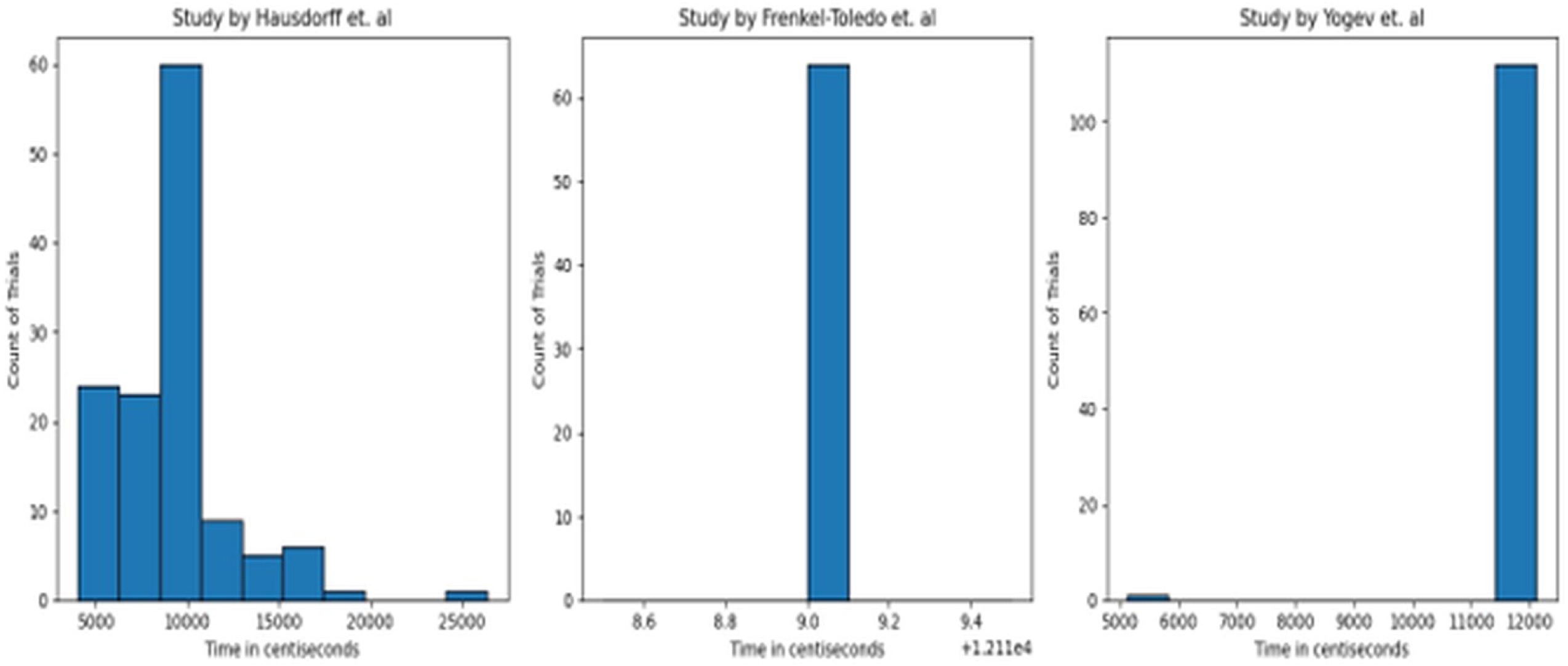

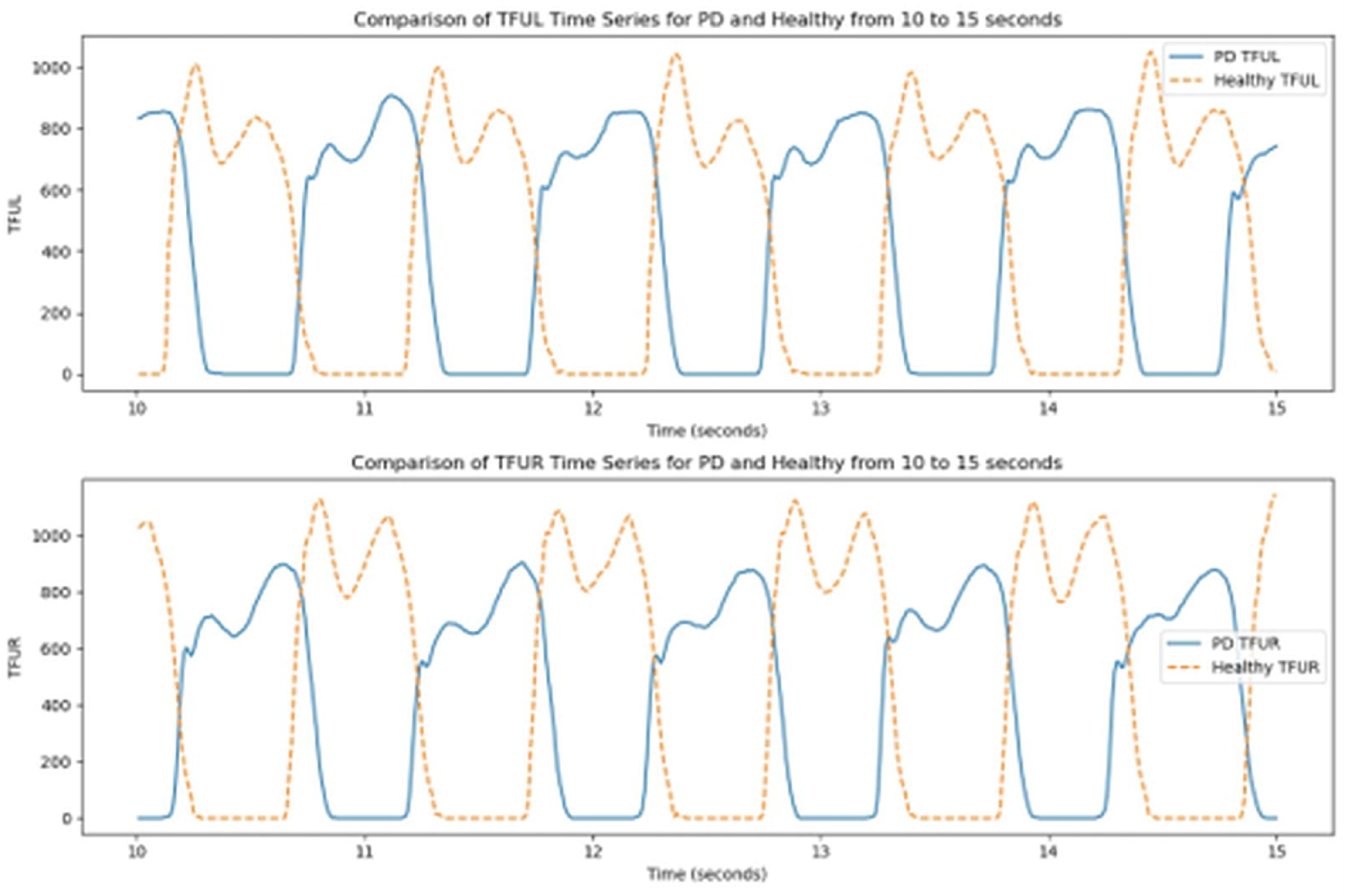

**Table 1:** Description of attributes in the utilized dataset. (Source: Yogev et al.(17))

|  |  |
| --- | --- |
| Attribute 1<sup>st</sup> | Time (in centiseconds) |
| Attributes 2<sup>nd</sup>-9<sup>th</sup> (Left Foot) | VGRF (in Newton) measurements for each of eight sensors. |
| Attributes 10<sup>th</sup>-17<sup>th</sup> (Right Foot) | VGRF measurements for each of the eight sensors. |
| Attribute 18<sup>th</sup> | Aggregated force under the left foot |
| Attribute 19<sup>th</sup> | Aggregated force under the right foot. |

\*\*\*Figure 3\*\*\* \*\*\*Figure 4 \*\*\*
\*\*\*Figure 5\*\*\*
\*\*\*Figure 6\*\*\*

### Data Preprocessing

Demographic information was used to stratify the data according to sex. This methodology was selected because recent studies have shown that VGRF-based gait analysis can effectively distinguish between Parkinson’s disease patients and healthy controls with considerable accuracy (Navita et al.(3)); however, the particular influences of age and sex stratification on diagnostic efficacy using this technique are still inadequately investigated. Research by Haaxma et al.(20) reveals that females exhibit the onset of Parkinson’s disease symptoms approximately 2.1 years later than males, with average onsets occurring at 53.4 years for females and 51.3 years for males. The preprocessing pipeline integrates domain-specific factors to improve clinical relevance and model generalizability. Individuals under 50 years of age (n = 3) were excluded to conform to the study’s emphasis on older populations exceeding 50 years, as PD gait patterns in younger groups may indicate atypical etiologies. The variables "total force under left foot" and "total force under right foot" were eliminated because they were derived from the summation of the sensor readings for each foot individually. This removal also facilitated a direct pipeline for transmitting the VGRF values from each of the sixteen sensors to the model for efficient real-time prediction. The variable identifying the time in centi-second was also removed. Although existing research emphasizes longer durations of 10 s and initial trims of 20 s (Navita et al.(3)), we implemented a 5 s window with a 10 s gait initiation trim, achieving a balance between temporal resolution and sample size sufficiency while reducing early stride variability. To standardize walking conditions, only the initial trial of each subject’s single-task walking was retained, omitting dual-task trials that introduced confounding cognitive loads(21). This targeted preprocessing adheres to established protocols for isolating PD-specific gait impairments during controlled clinical observation. These modifications highlight the essential importance of task-specific data curation in deep learning applications, where improper segmentation or the inclusion of diverse tasks can artificially enhance training performance at the expense of real-world validity.

### Proposed Models

#### LSTM Network

A Long Short-Term Memory (LSTM) network mitigates the vanishing and exploding gradient issues inherent in traditional Recurrent Neural Networks (RNN) by utilizing memory cells and gated mechanisms (input, forget, and output gates) to preserve essential temporal dependencies, as introduced by Hochreiter et al(22). As the name suggests, the forget gate determines which elements of information from the preceding cell state should be retained and which should be removed. This is accomplished by employing a sigmoid activation function on the concatenation of the preceding hidden state and the current input, producing values ranging from 0 to 1, which acts as a filter for the prior cell state.

After determining what to forget, the input gate reviews the present input and produces new candidate values for potential inclusion in the cell state. Sigmoid activation identifies the elements that should be updated, whereas a separate layer constructs a vector of potential values to be incorporated. The next step involves updating the cell state by merging the preserved information from the previous state filtered through the forget gate with the newly generated candidate values scaled by the input gate.

The output gate utilizes the revised cell state to determine the information to be transmitted as a hidden state, which is subsequently employed in further computations and layers. This mechanism allows the LSTM network to balance the retention of long-term memory with the integration of new inputs, rendering it particularly effective for tasks such as distinguishing between PD and healthy individuals. Equations 1 through 6 in Table 2 elucidate the processes occurring within an LSTM cell, as illustrated in Figure 7.

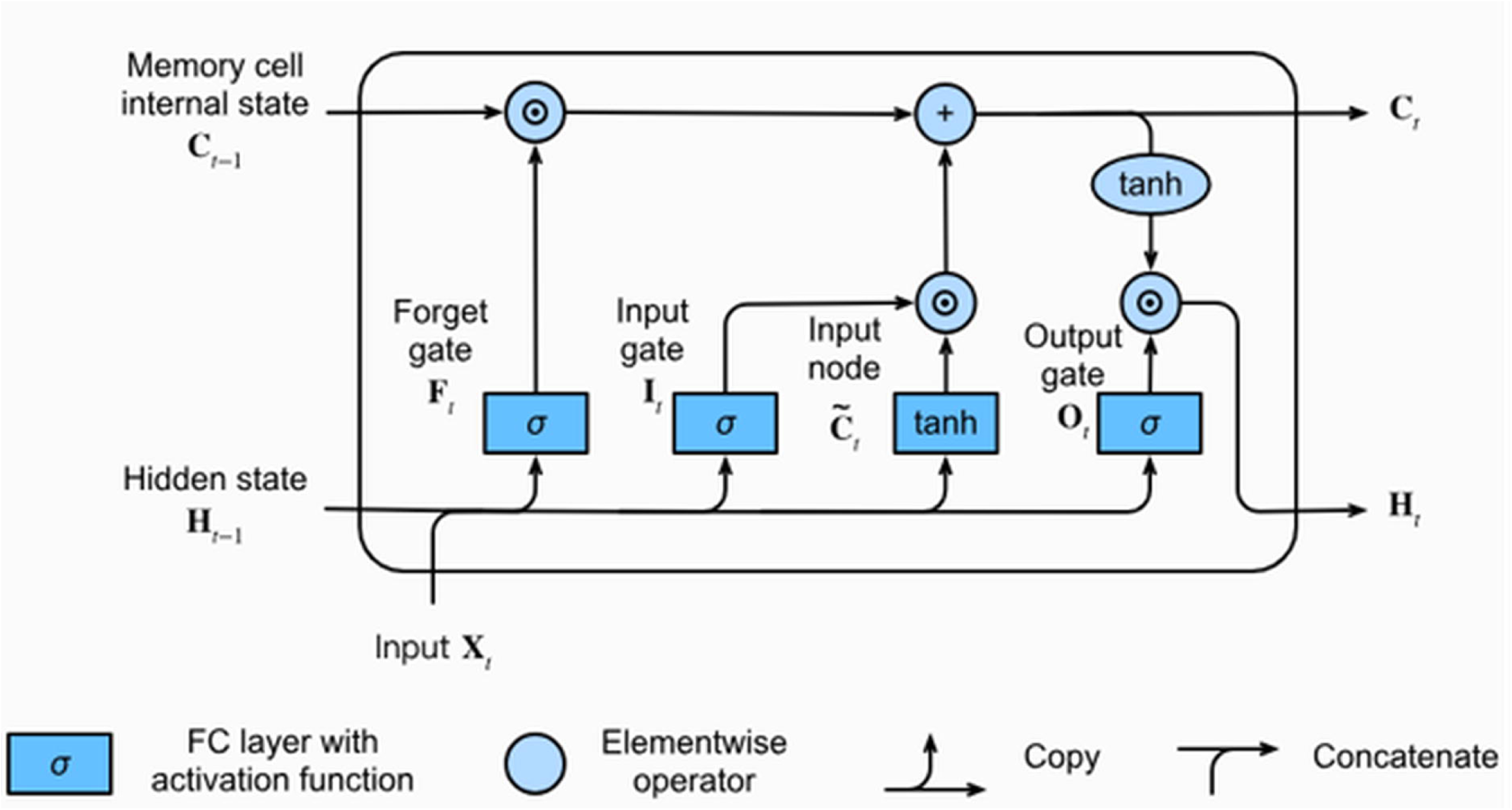

**Table 2:**
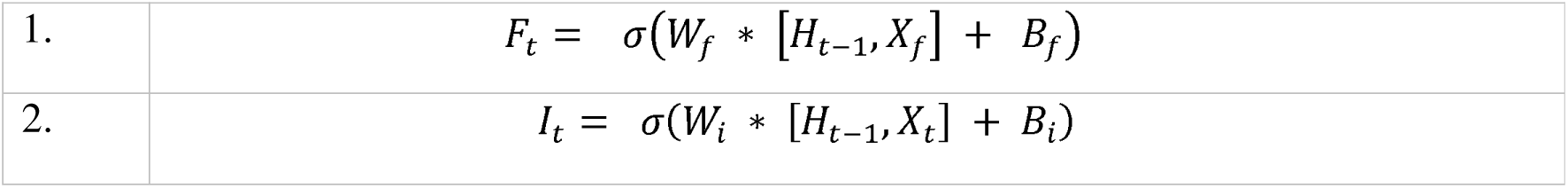
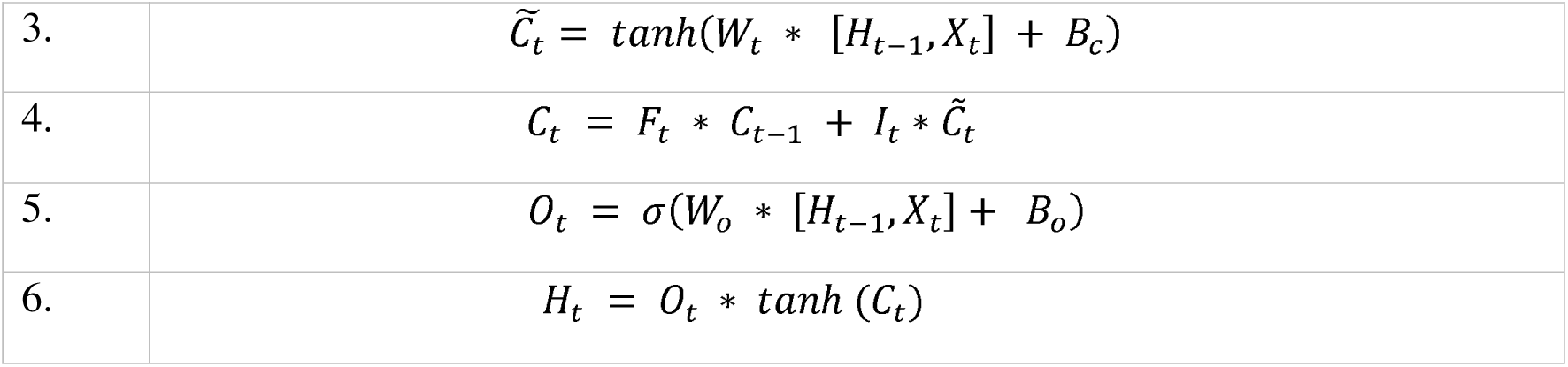
LSTM mathematical equations. (Source: https://zhuanlan.zhihu.com/p/39624477)

#### Bidirectional LSTM Network

Bidirectional LSTM networks, initially presented by Graves et al.(23), augment unidirectional LSTM networks by incorporating a parallel LSTM layer capable of capturing temporal patterns and dependencies in the reverse direction. They can process input sequences in both forward and backward directions, rendering them an effective model for signal processing and modeling, where capturing context from both the past and the future is crucial. They require increased training duration and computational resources relative to the LSTM model. Figure 8 illustrates its operation.

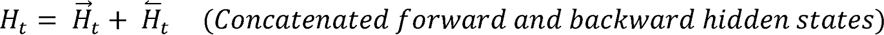

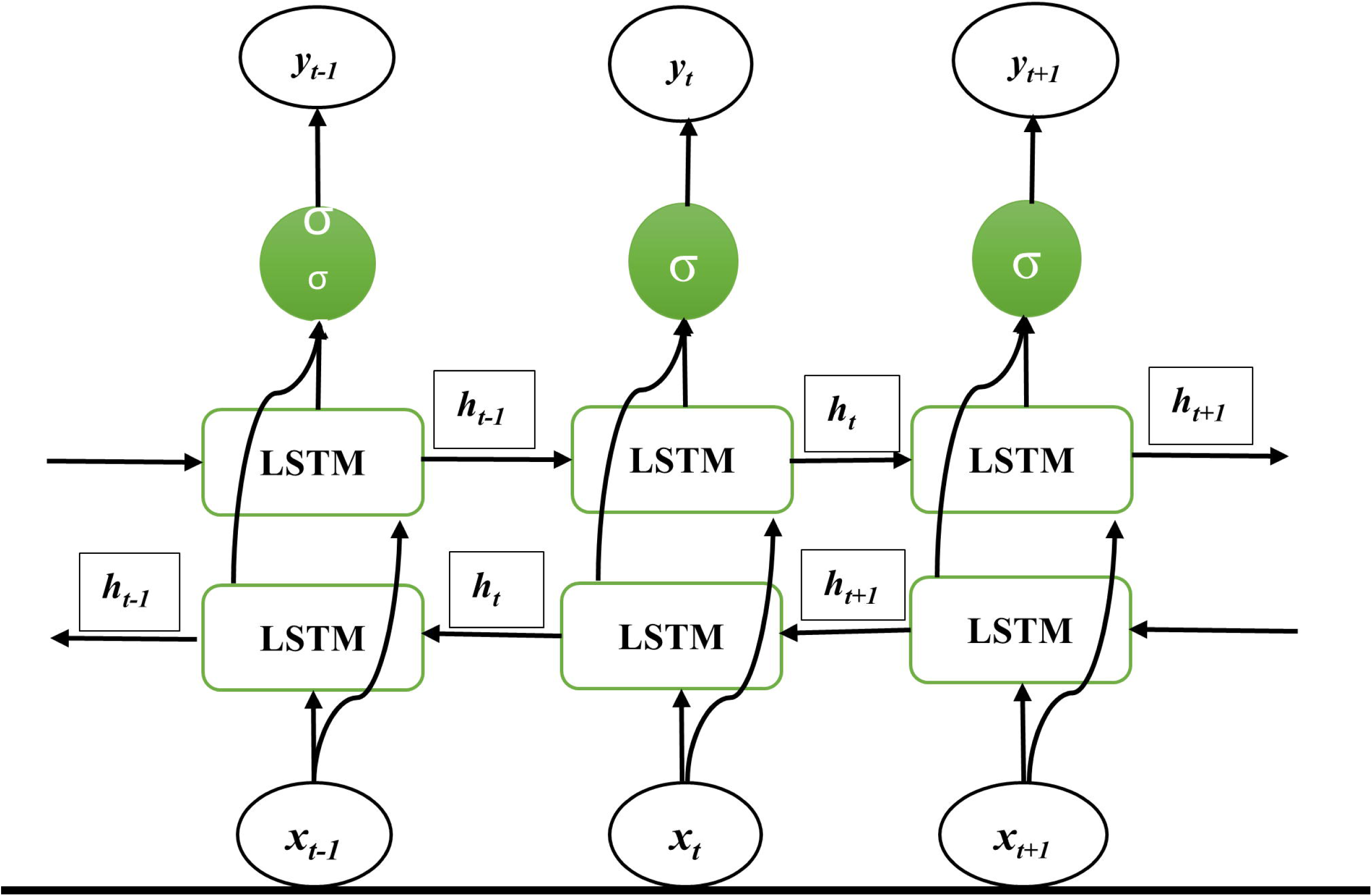

#### Gated Recurrent Unit Network

Gated Recurrent Unit (GRU) networks were introduced by Cho et al.(24) and are specifically engineered to manage sequential data by comprehending and retaining intricate patterns. Their components include gates and memory units that enable the GRU network to understand relationships across various time intervals, providing improved computational efficiency relative to traditional LSTM networks. It comprises two gates: a reset gate and an update gate. The reset gate ascertains the extent to which the prior hidden state should be disregarded, whereas the update gate evaluates the degree to which the current input should be utilized to update the hidden state. Both gates are associated with the hidden state. They differ from LSTMs by merging the forget and input gates into a singular "update gate," which facilitates the precise regulation of information flow to effectively manage essential data within sequences. The GRU network lacks an output gate and utilizes a more concise set of parameters than the LSTM network. Yang et al.(25) examined the dual aspects of performance and computational cost, revealing that the performance-cost ratio of GRU surpassed that of LSTM. Figure 9 illustrates the architecture of a GRU cell, and the accompanying equations elucidate the computations occurring within it.

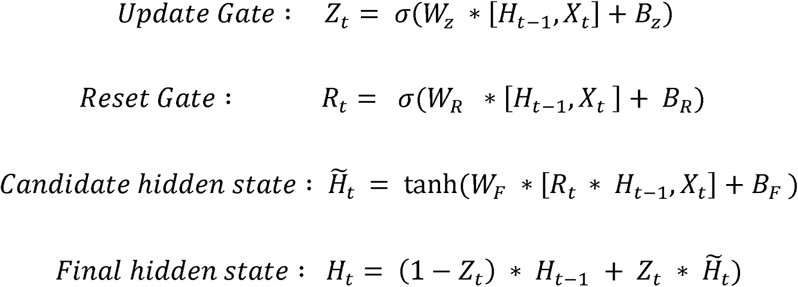

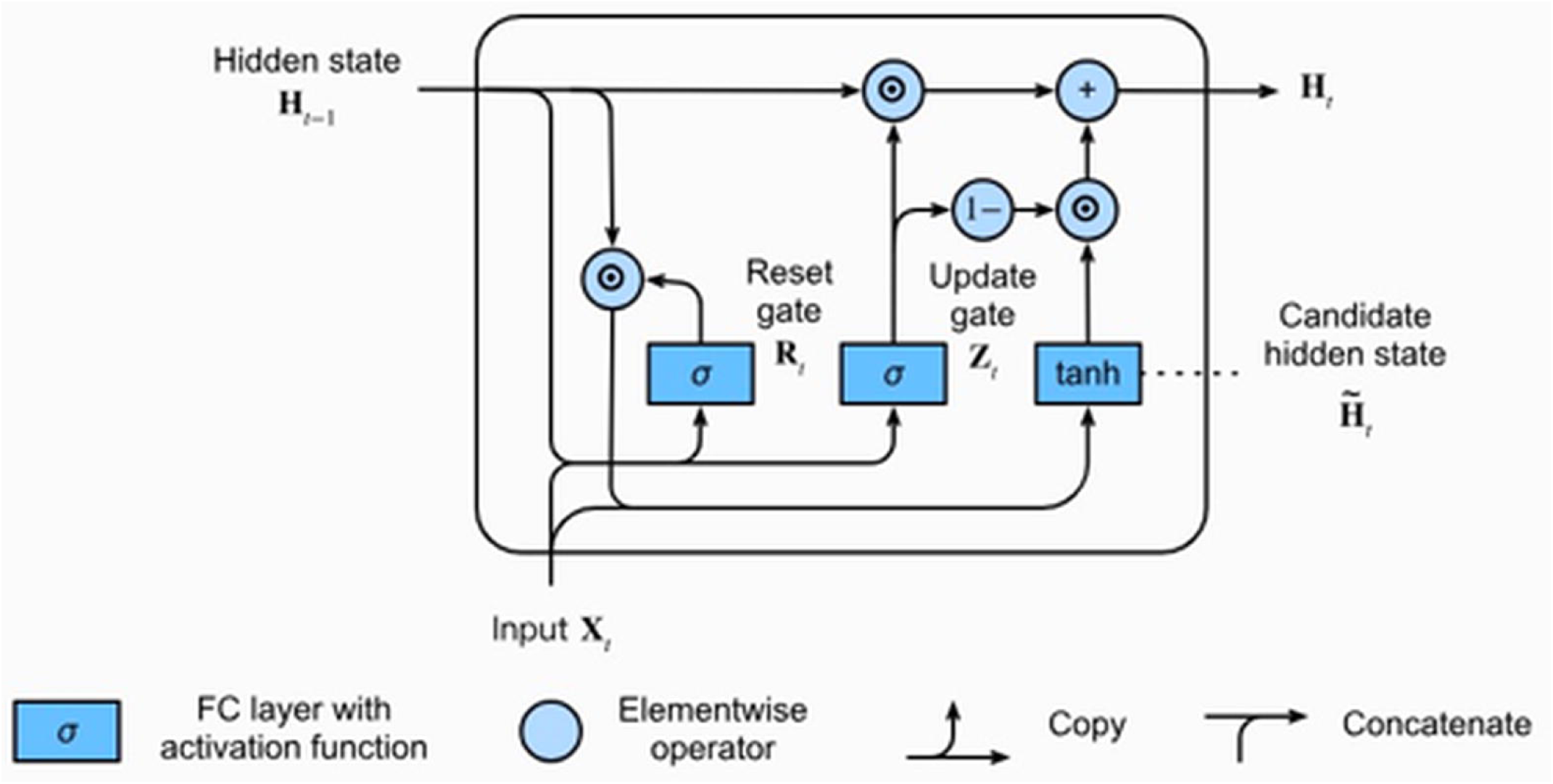

#### Bidirectional GRU network

Bidirectional Gated Recurrent Unit (bi-GRU) networks enhance the functionality of unidirectional GRU architectures by integrating two parallel GRU layers: one that processes input sequences in the forward temporal direction and another that processes them in reverse. This dual processing allows the model to capture contextual dependencies from both prior and subsequent elements of sequential data, such as VGRF signals in the Parkinsonian gait analysis. In contrast to unidirectional models, the bidirectional architecture of bi-GRU is particularly proficient in situations that require a comprehensive temporal context, such as detecting stride irregularities or gait initiation anomalies. Despite being computationally more demanding than conventional GRUs owing to the increased parameterization, the superior feature extraction capabilities of bi-GRUs validate their application in clinical motion analytics. Figure 10 provides a visual representation that resembles the structure of the bidirectional LSTM network.

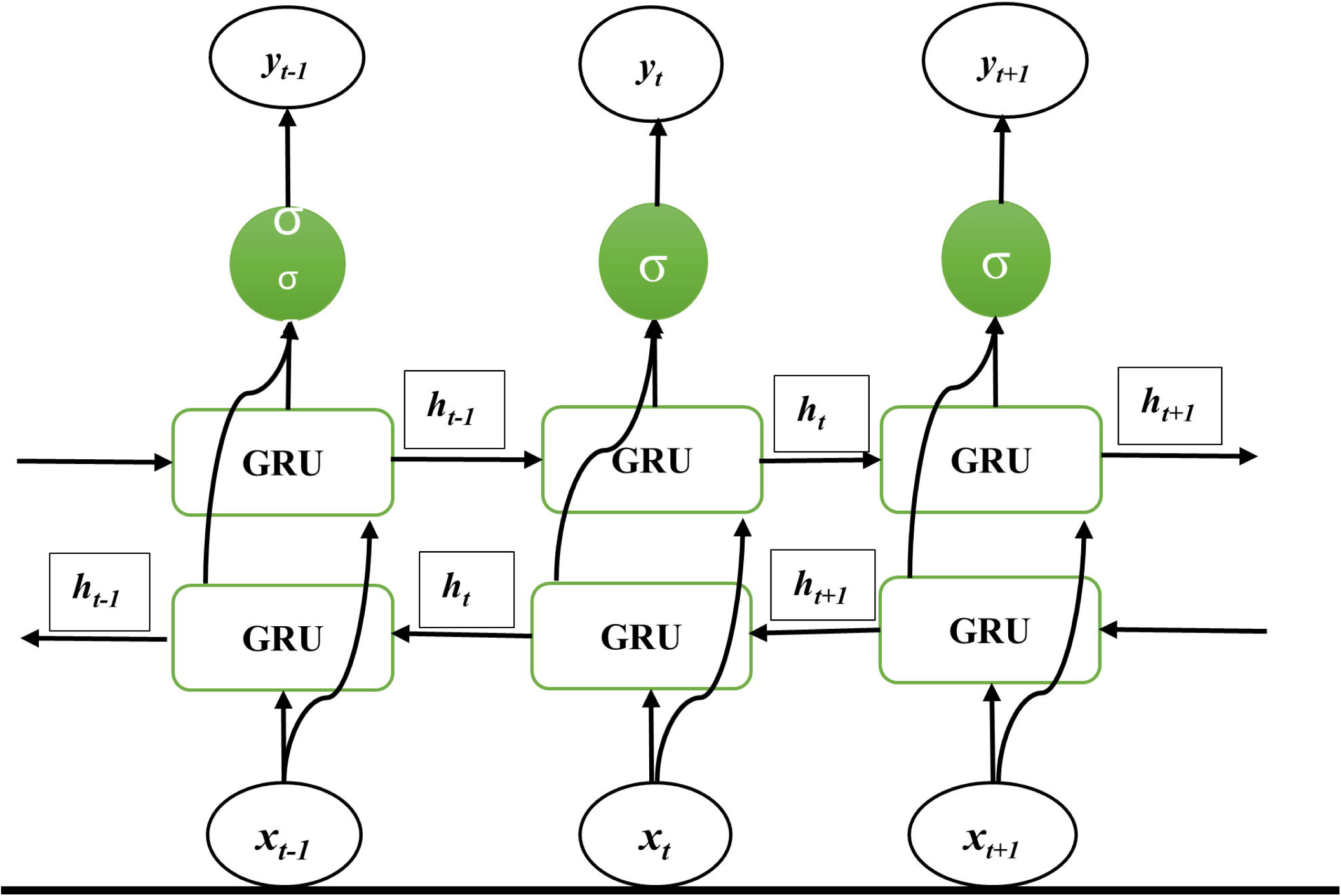

### Model Development

The suggested DL architecture for detecting PD using gait-based VGRF sensor data maintains a uniform structural framework across all four models, differing solely in the selection of optimal hyperparameters, as illustrated in Figure 11.

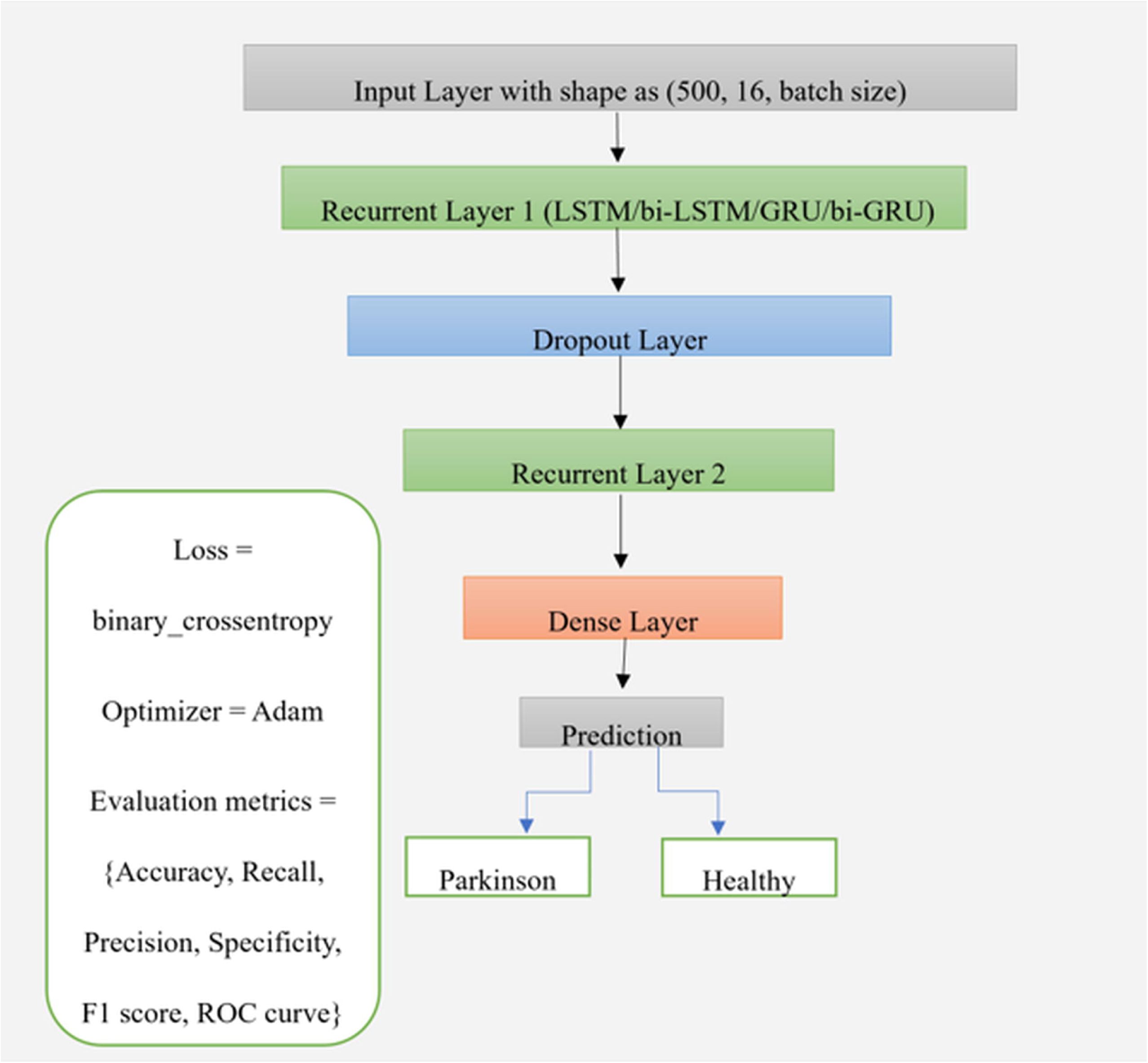

#### L2 regularization

L2 regularization is a commonly employed method in deep learning to mitigate the effects of overfitting and produce enhanced model generalization. This is achieved by incorporating a penalty term into the original loss function. This penalty deters excessive weight values, thereby streamlining the model and reducing its susceptibility to noise in training data. Mir et al.(7) also illustrated the efficacy of this regularization method in their model development. An identical methodology was selected for model training based on their work. The mathematical expression is as follows:

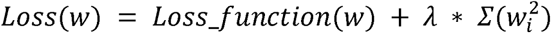

The "loss_function" refers to the binary cross-entropy loss, "w" denotes the model weights acquired during an iteration, "w_i_" signifies individual coefficients, and "λ" (lambda or alpha) is the regularisation parameter that regulates the intensity of the penalty. An elevated lambda value leads to enhanced regularization, promoting reduced non-zero coefficients and preventing any individual feature from dominating the model’s predictions.

#### Adam Optimizer

An optimization technique extensively employed in training deep neural networks, yielding faster convergence rates than traditional optimization algorithms, was introduced by Kingma et al.(26) by integrating the characteristics of RMSprop and momentum-based gradient descent methods. Mir et al.(7) utilized it in their model development to attain remarkable convergence speed and predictive accuracy regarding the freezing of gait. Rehman et al.(27) also demonstrated its efficacy in training deep neural networks for the detection of Parkinson’s disease. Its capacity to adjust to the calculated gradients of the loss function and iteratively modify learning rates renders it resilient to noisy gradients while simultaneously ensuring efficient memory usage and expedited convergence through hyperparameter selection.

#### Dropout Layer

A regularization technique for deep neural networks, introduced by Srivastava et al., that alters intermediate neural connections by randomly deactivating incoming neurons and their connections with a probability of *p*. This strategy prevents hidden units from becoming overly reliant on one another (co-adaptation), promoting stronger and more generalized learning while mitigating overfitting. Mir et al.(7), Rehman et al.(27), and additional studies employing DL models extensively utilize this technique to enhance model robustness and generalizability. We established the value *p* for each model through hyperband hyperparameter tuning.

#### Early Stopping

A distinct regularization technique that halts the training process upon detecting overfitting by assessing the model’s performance on the validation set throughout the training phase. In 1998, Prechelt(28) emphasized its effectiveness, a period of time when computational resources of time and space were constrained. Currently, it enhances the training process by halting it based on performance metrics, specifically "val_accuracy," and the "patience" parameter, which determines the number of epochs to wait before assessing whether learning has improved or declined. It also enabled us to recover the optimal weights acquired when the model attained peak performance during training. This led to employing a reduced number of epochs during the training phase compared to the initially defined range based on the study by Mir et al.(7)

#### Hyperparameter Tuning

Hyperparameters are model parameters not directly acquired during training; instead, they are configuration variables that govern the model’s learning process. They are established prior to training and remain unchanged throughout the training process. Hyperparameter tuning is crucial for the network’s learning of the underlying function within an extensive hyperparameter search space. Hyperband hyperparameter tuning reallocates resources dynamically based on intermediate outcomes, rendering it exceptionally efficient for iterative algorithms such as deep learning models, as demonstrated by Li et al.(29). Our proposed models employed a consistent methodology to guarantee the efficient learning of optimal hyperparameter configurations for each individual model, as presented in Table 3.

**Table 3:** Modeling Hyperparameters for Males and Females.

| **Model** | *Units in 1<sup>st</sup> Recurrent Layer* | *L2 regularization value* | *Dropout rate* | *Units in 2<sup>nd</sup> Recurrent Layer* | *Learning Rate for Adam Optimizer* | *Total epochs of training with early stopping* |
| --- | --- | --- | --- | --- | --- | --- |
| **LSTM (Males)** | 256 | 0.0005 | 0.3 | 96 | 0.0005 | 80 |
| **Bi-LSTM (Males)** | 128 | 0.0001 | 0.1 | 64 | 0.0005 | 80 |
| **GRU (Males)** | 256 | 0.0005 | 0.1 | 64 | 0.001 | 80 |
| **Bi-GRU (Males)** | 256 | 0.0005 | 0.1 | 32 | 0.001 | 50 |
| **LSTM (Females)** | 256 | 0.005 | 0.1 | 64 | 0.0005 | 64 |
| **Bi-LSTM (Females)** | 256 | 0.0005 | 0.2 | 64 | 0.001 | 30 |
| **GRU**<br>**(Females)** | 256 | 0.005 | 0.2 | 128 | 0.001 | 27 |
| **Bi-GRU**<br>**(Females)** | 256 | 0.0005 | 0.2 | 32 | 0.005 | 43 |

### Evaluation Metrics

Confusion matrices were created to assess the performance of the developed models. Performance metrics such as accuracy, recall, precision, and F1 scores were computed. This aligns with research conducted by scholars such as Navita et al.(3), Mir et al.(7), Rehman et al.(27) Pham(30), Rashnu et al.(31), and others, wherein PD detection was approached as a classification issue. Accuracy as a performance metric quantifies the ratio of correct predictions (true positives and true negatives) for both classes relative to the total predictions made by the model, thereby explaining the model’s efficacy for each class(27). The percentage of true positives to all actual positives is measured by recall(27), including false negatives, which involve misclassifying Parkinson’s disease patients as healthy, are crucial in clinical contexts, as undiagnosed Parkinson’s disease can postpone treatment and exacerbate outcomes. It guarantees the detection of the majority of Parkinson’s cases. Precision quantifies the ratio of true positives predicted by the model to the aggregate of both true positives and false positives. Misclassifying a healthy individual as a Parkinson’s patient may result in superfluous expenditure of resources, including time and money, as well as induce anxiety for the individual. Precision guarantees that the model reduces such errors. F1 scores amalgamate the performance metrics of recall and precision as their harmonic mean, thereby balancing both into a singular metric. This is especially beneficial for imbalanced datasets such as PD classification, where both false positives and false negatives carry substantial repercussions.

Another crucial evaluation tool is the Receiver Operating Characteristic curve, abbreviated as ROC, which assesses and helps strike an equilibrium between the true positive rate (sensitivity) and the false positive rate across various thresholds of a binary classifier(27), as per the tolerance criteria of misclassification. The measurement of the total differentiation between classes is achieved by calculating the area under the curve (AUC). ROC-AUC is robust against class imbalance and provides insights into the model’s efficacy in distinguishing between healthy individuals and those with Parkinson’s disease. It is particularly beneficial for model comparison or the optimization of decision thresholds aligned with therapeutic aims.

## Results

### Male subjects

The aforementioned performance metrics are presented herein. Figures 12 to 15 present confusion matrices for the comparison of models developed for males. Positive and negative class labels are denoted as “1” and “0,” respectively, with “1” indicating a PD subject and “0” indicating a healthy subject.

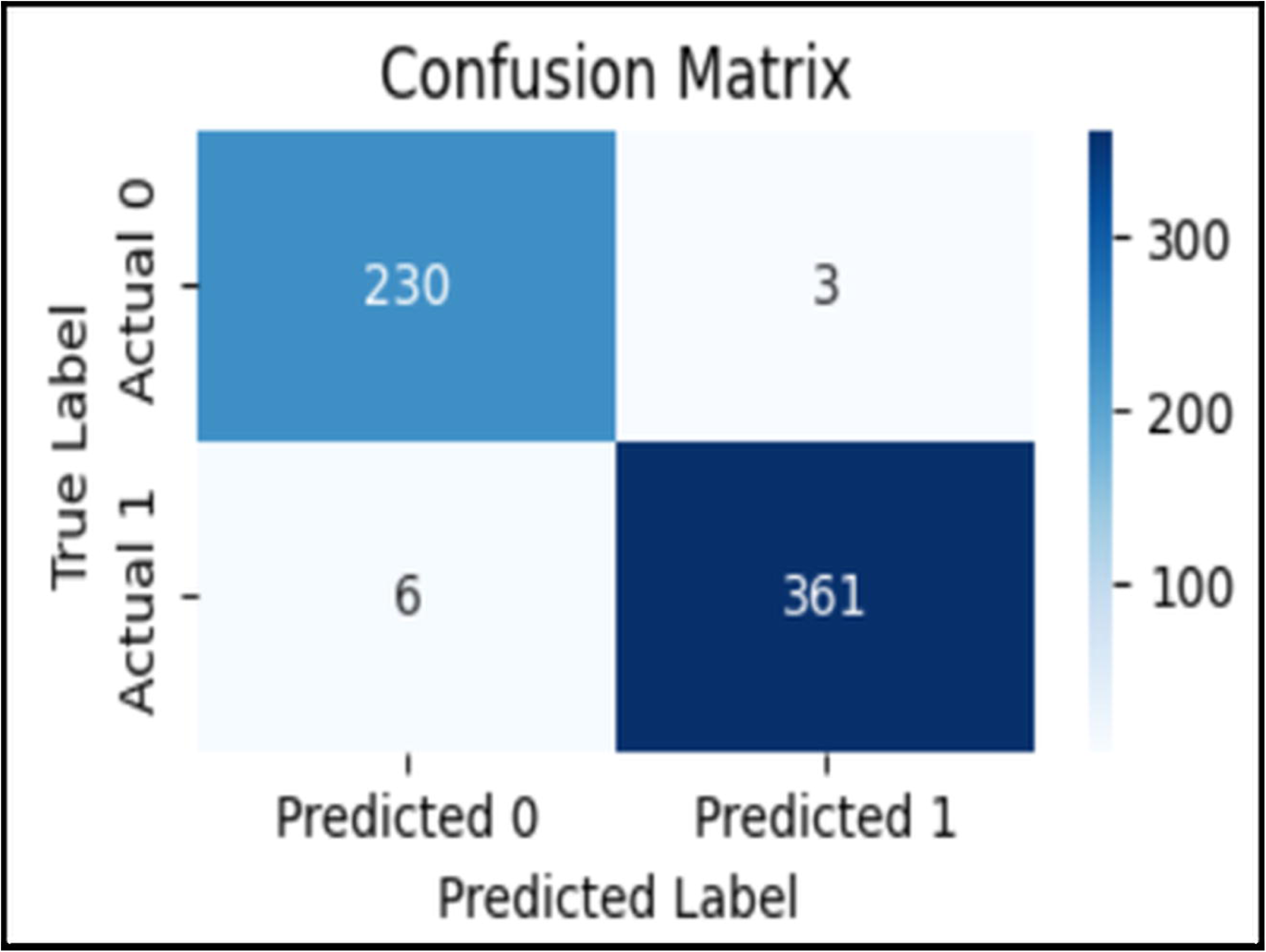

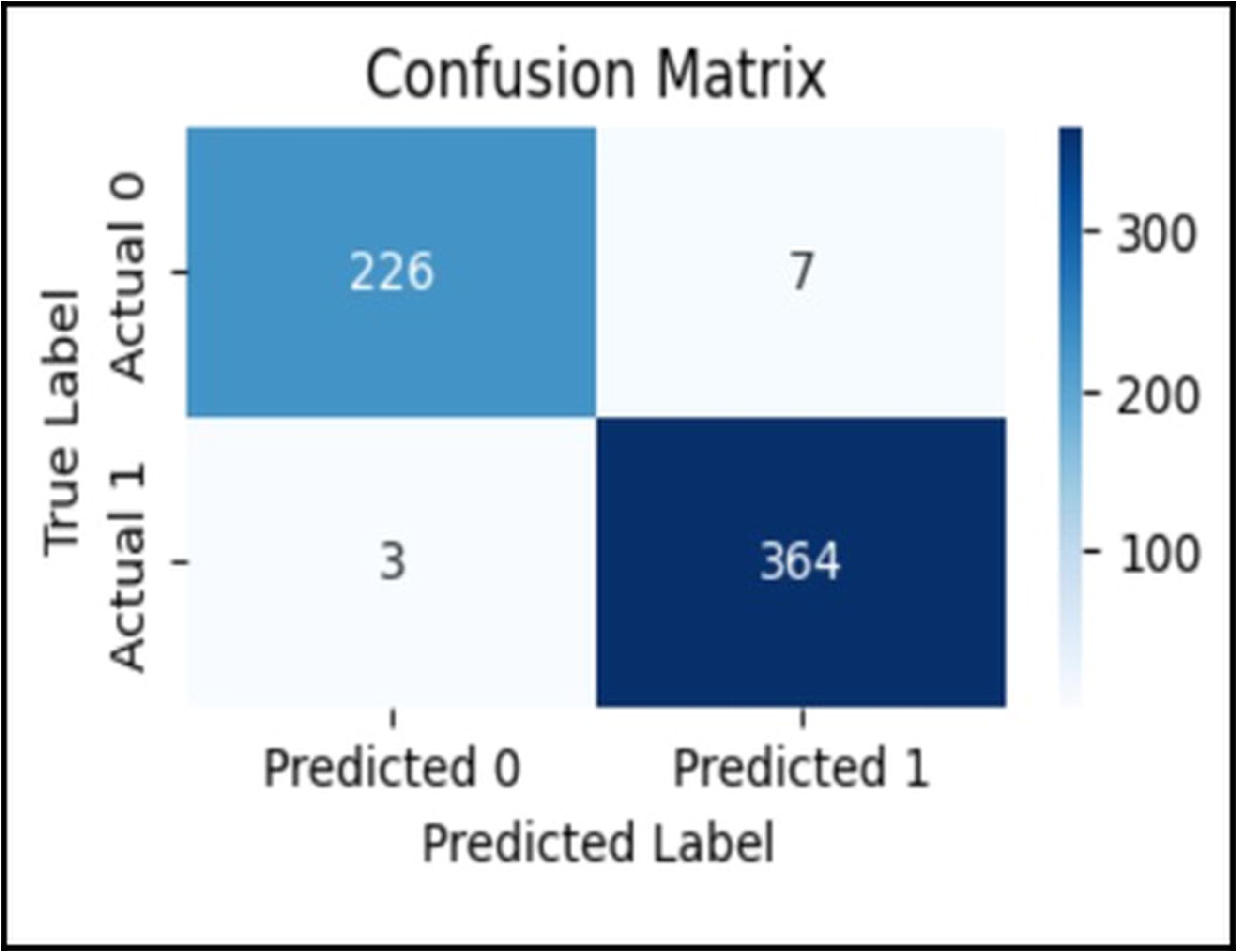

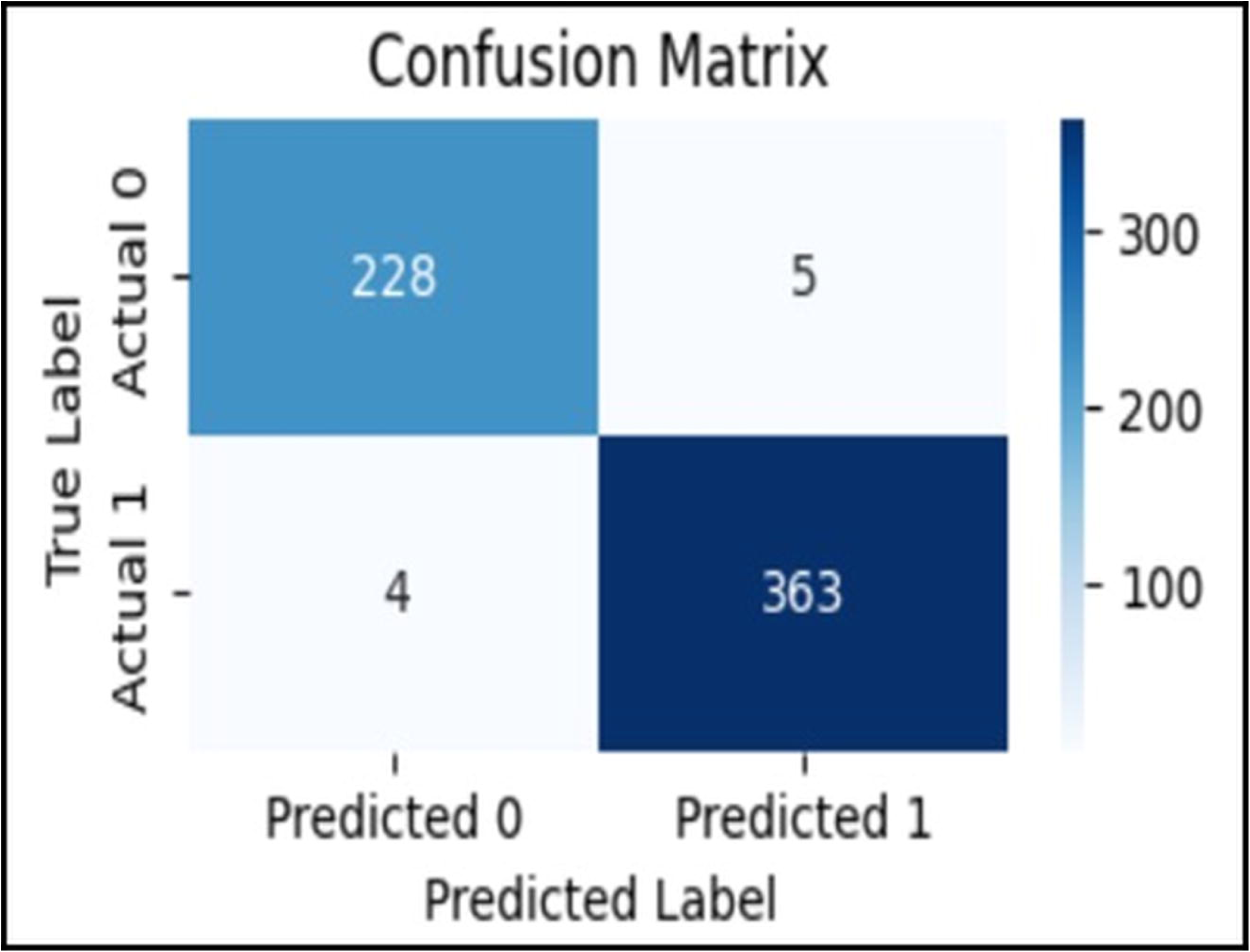

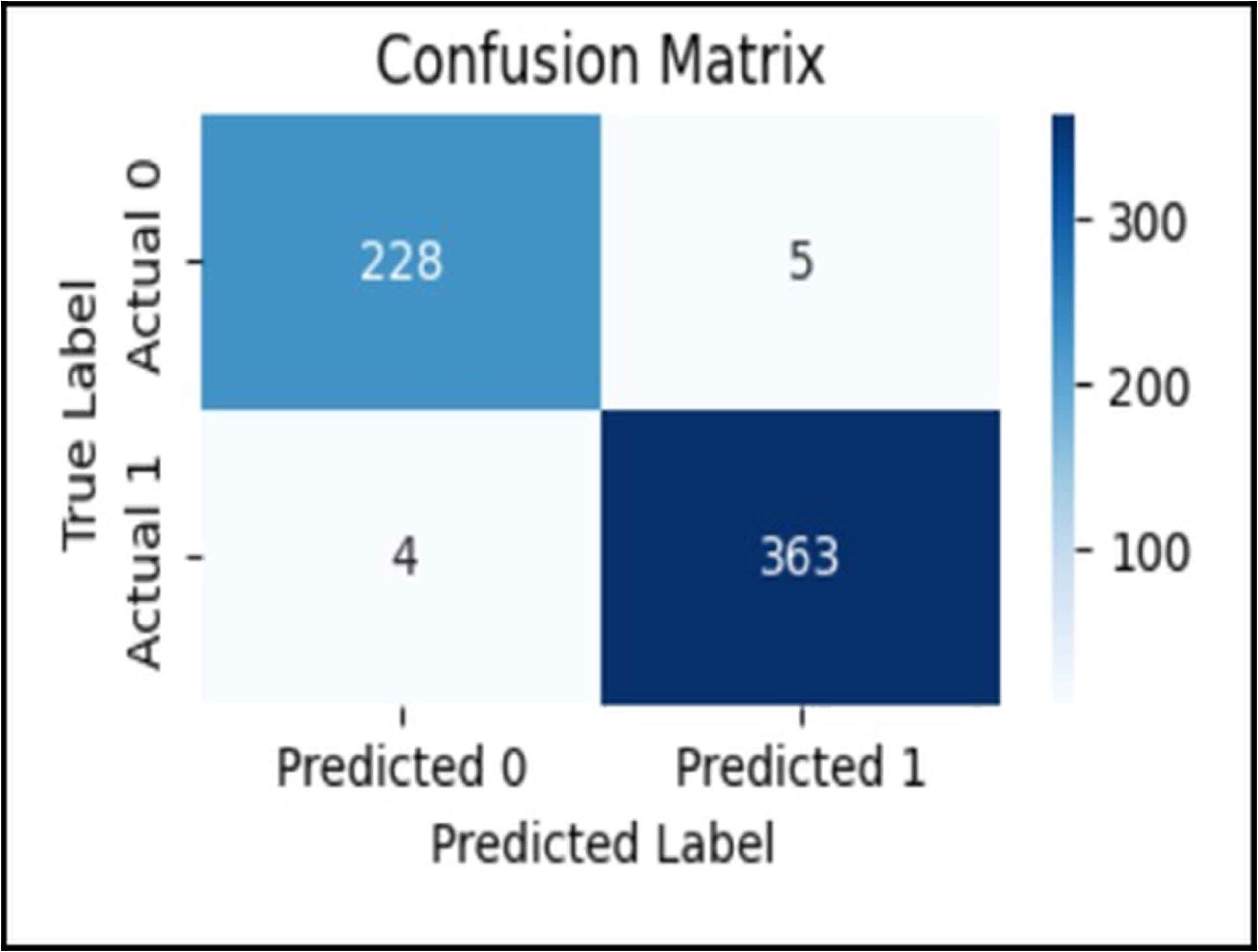

Figure 16 presents the individual performance metrics of the models.

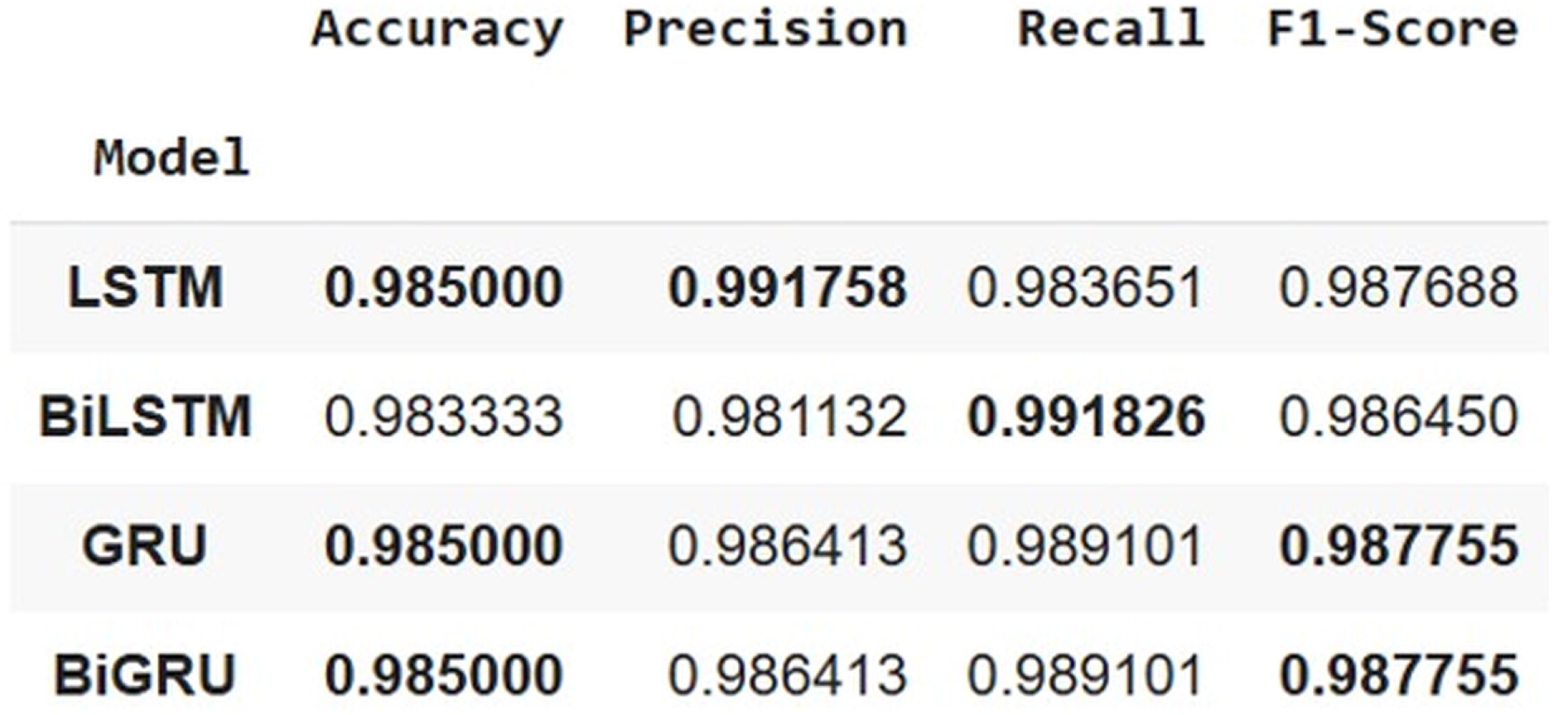

The ROC-AUC plot of the models is presented in Figure 17.

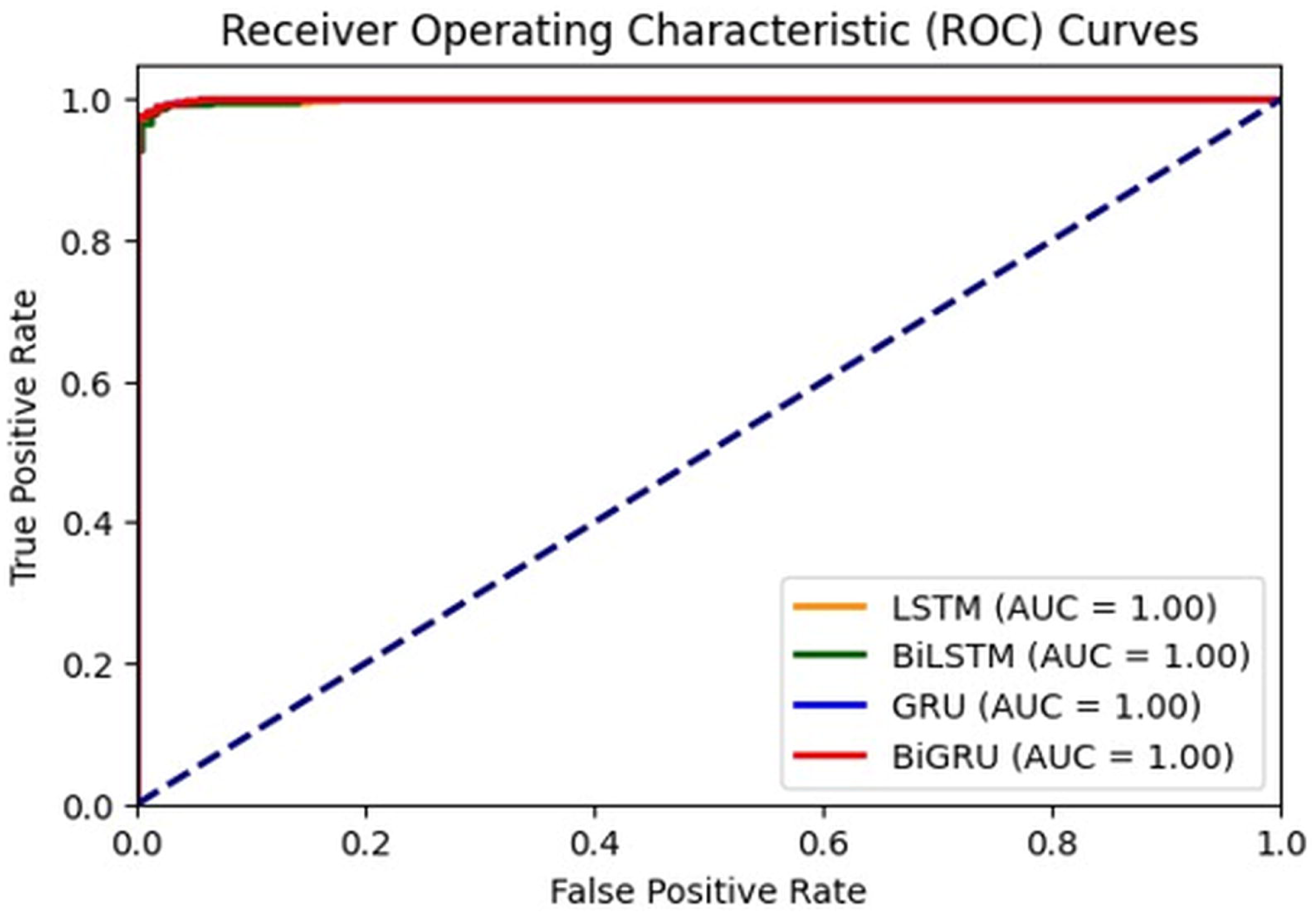

Since GRU models, whether unidirectional or bidirectional, demonstrate comparable performance to LSTM-based models, their reduced parameter requirements render them more efficient in training relative to LSTM-based models. The findings indicate that these models are equally proficient in identifying pertinent sequential patterns in the data for males and may be favored over alternative models.

### Female Subjects

The confusion matrices for each model are presented from Figure 18 to 21 for comparison.

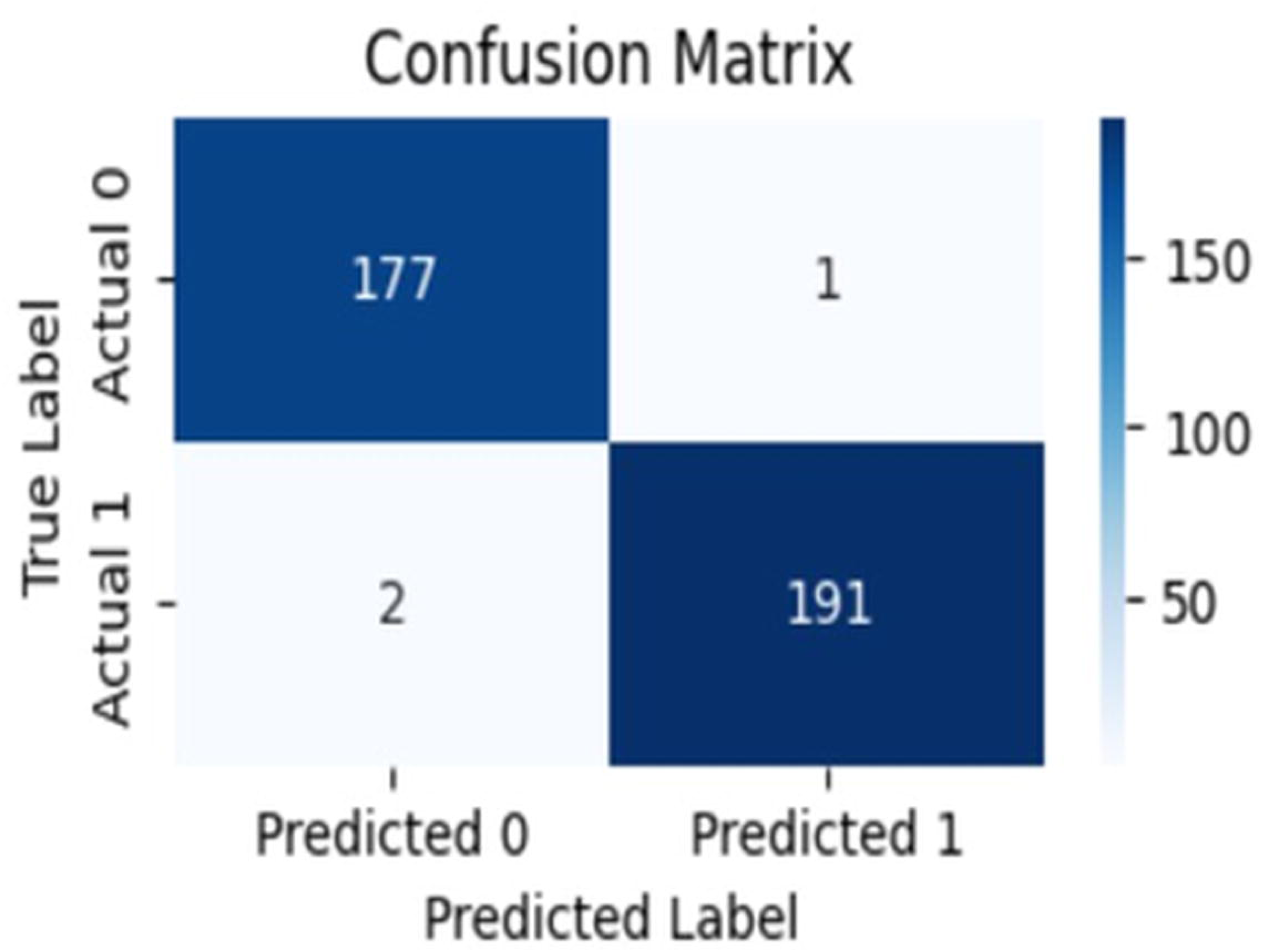

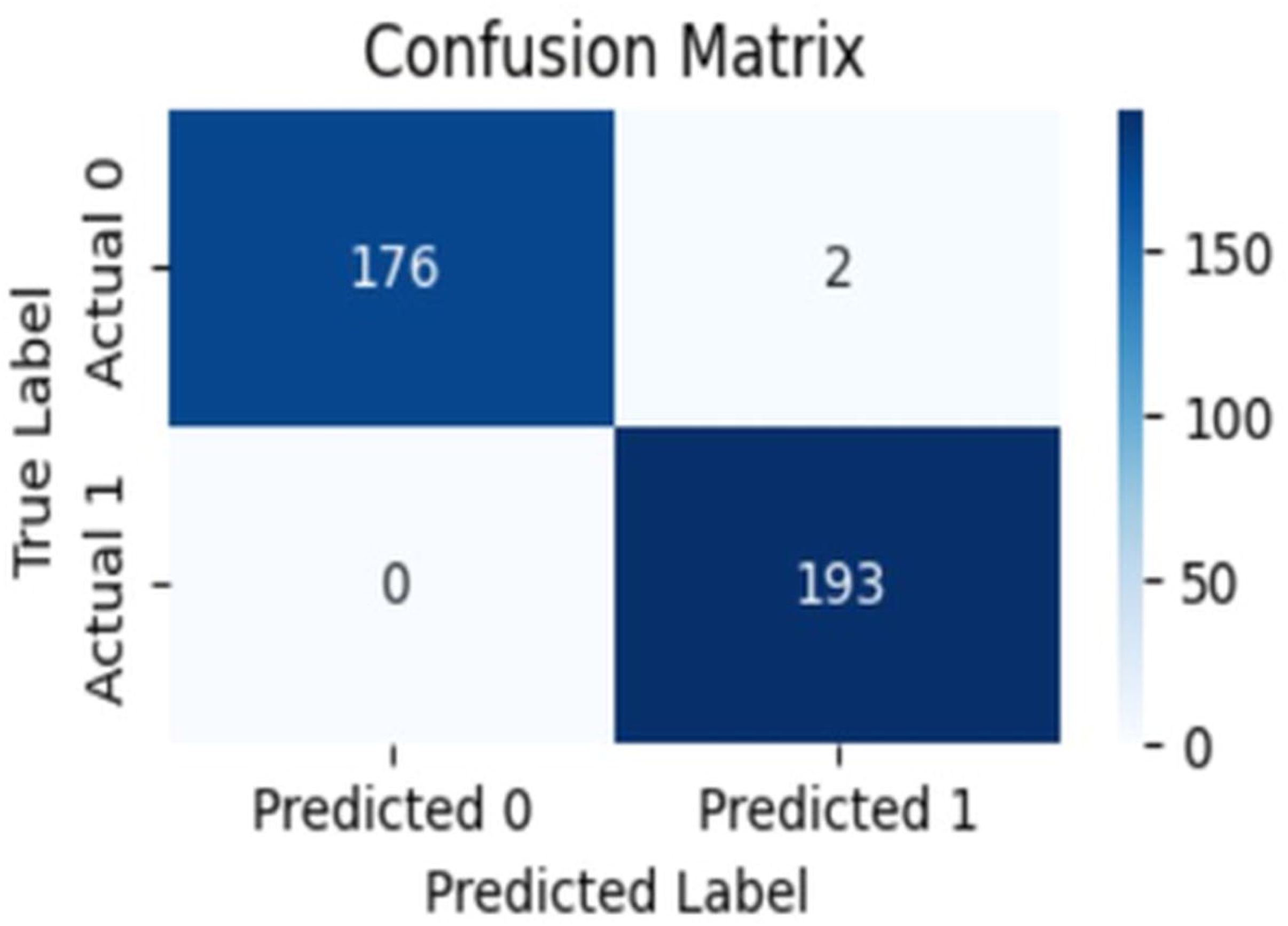

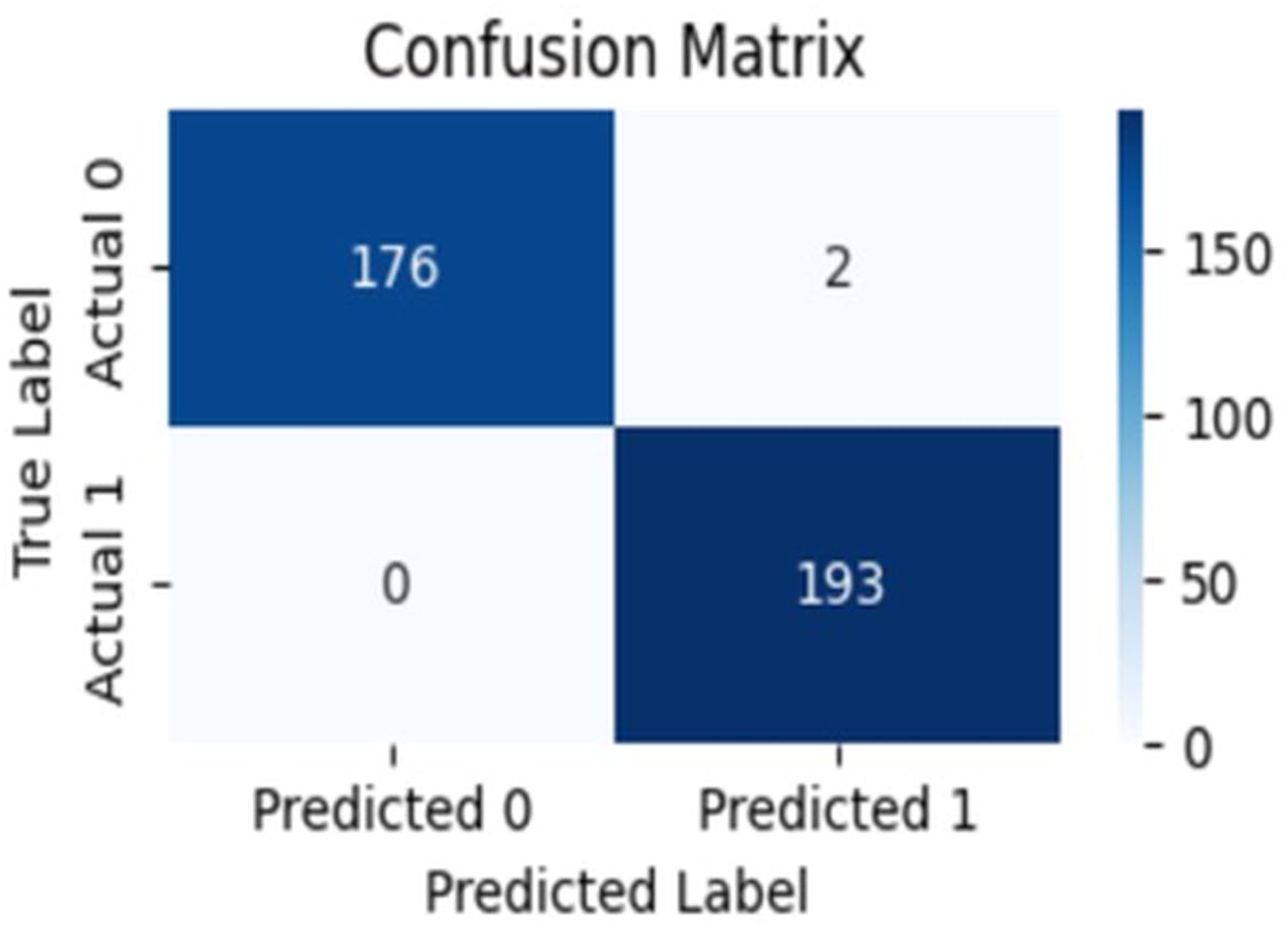

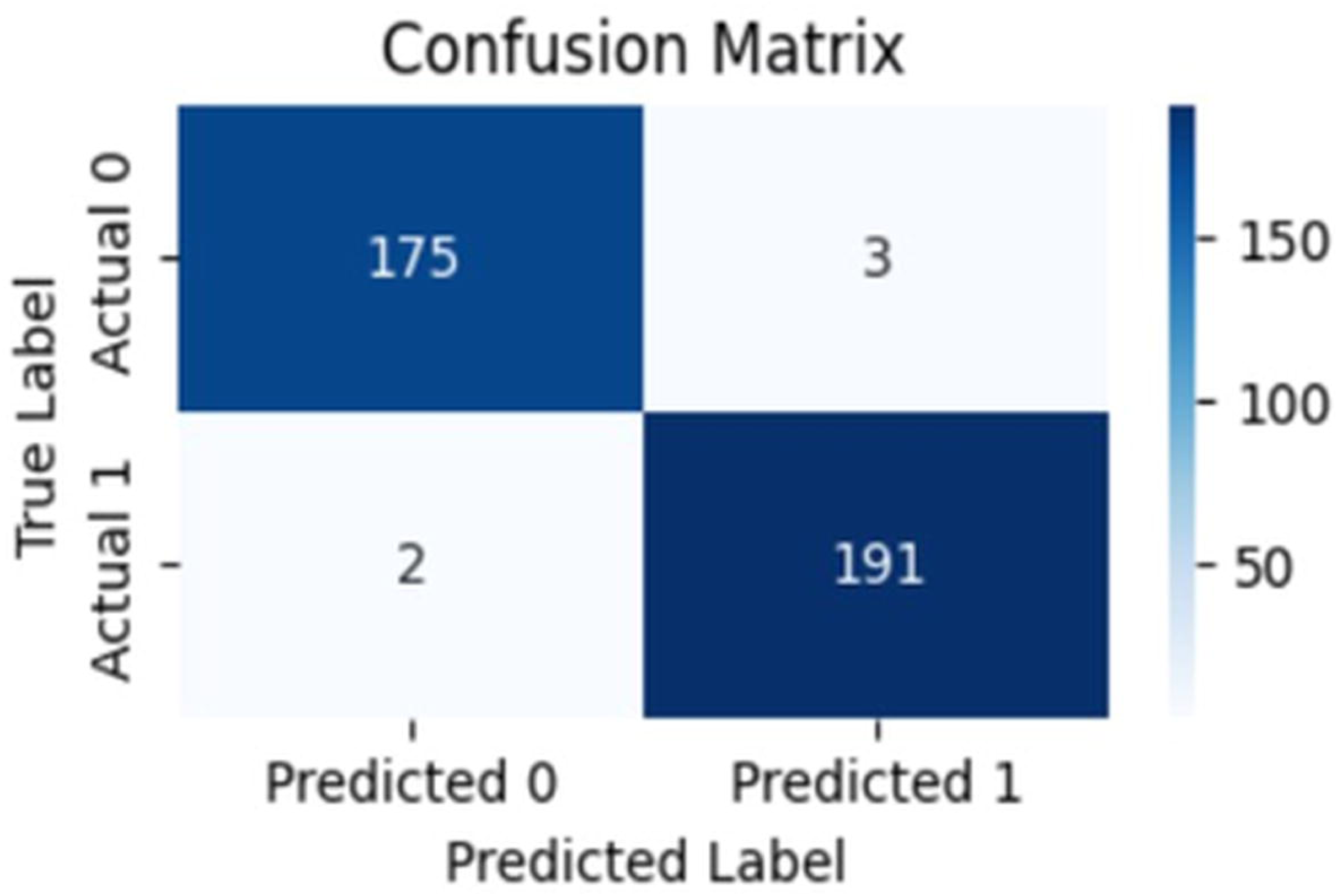

Individual performance metrics of models are highlighted in Figure 22.

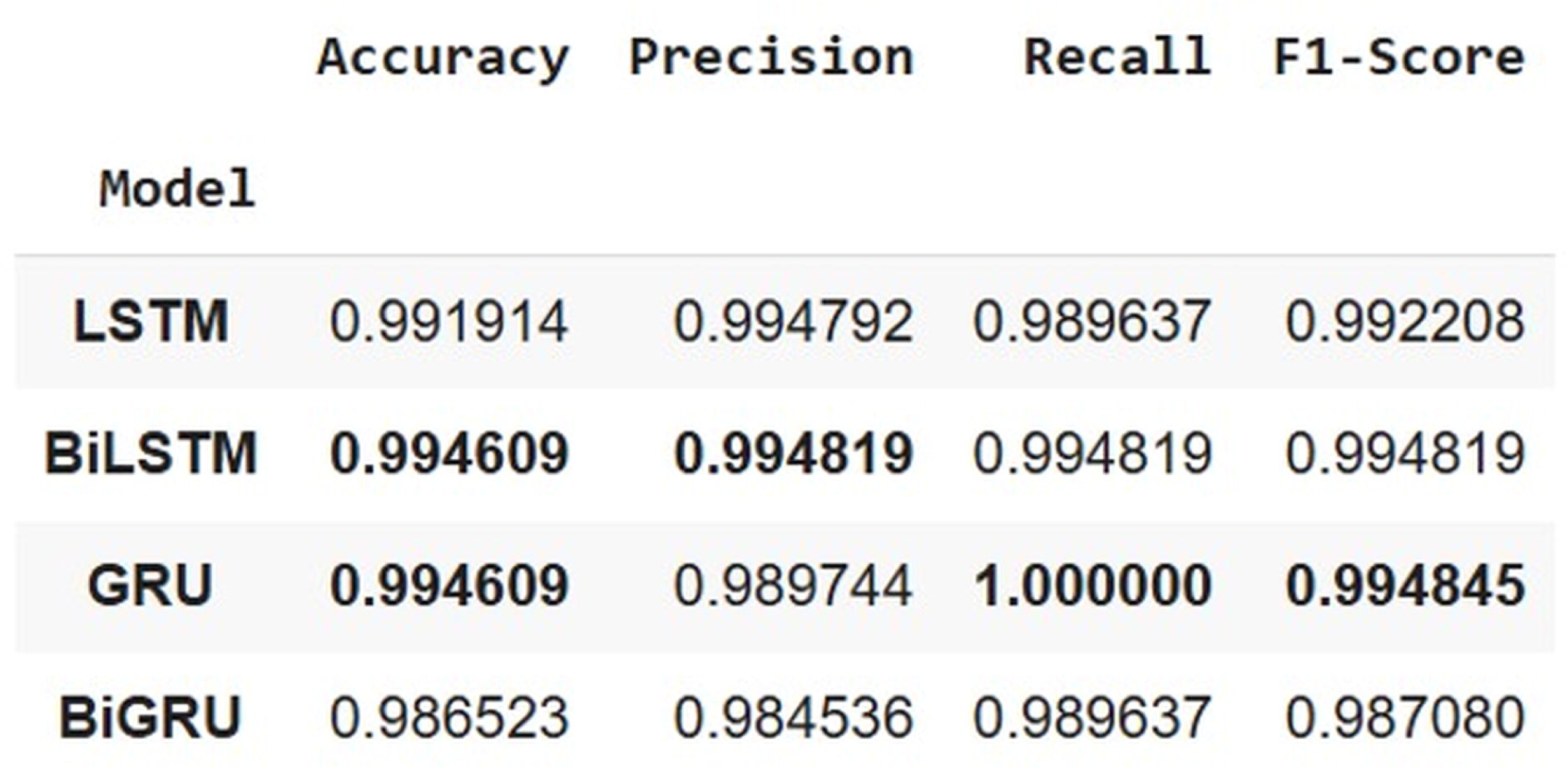

The ROC-AUC plot of the models is presented in Figure 23.

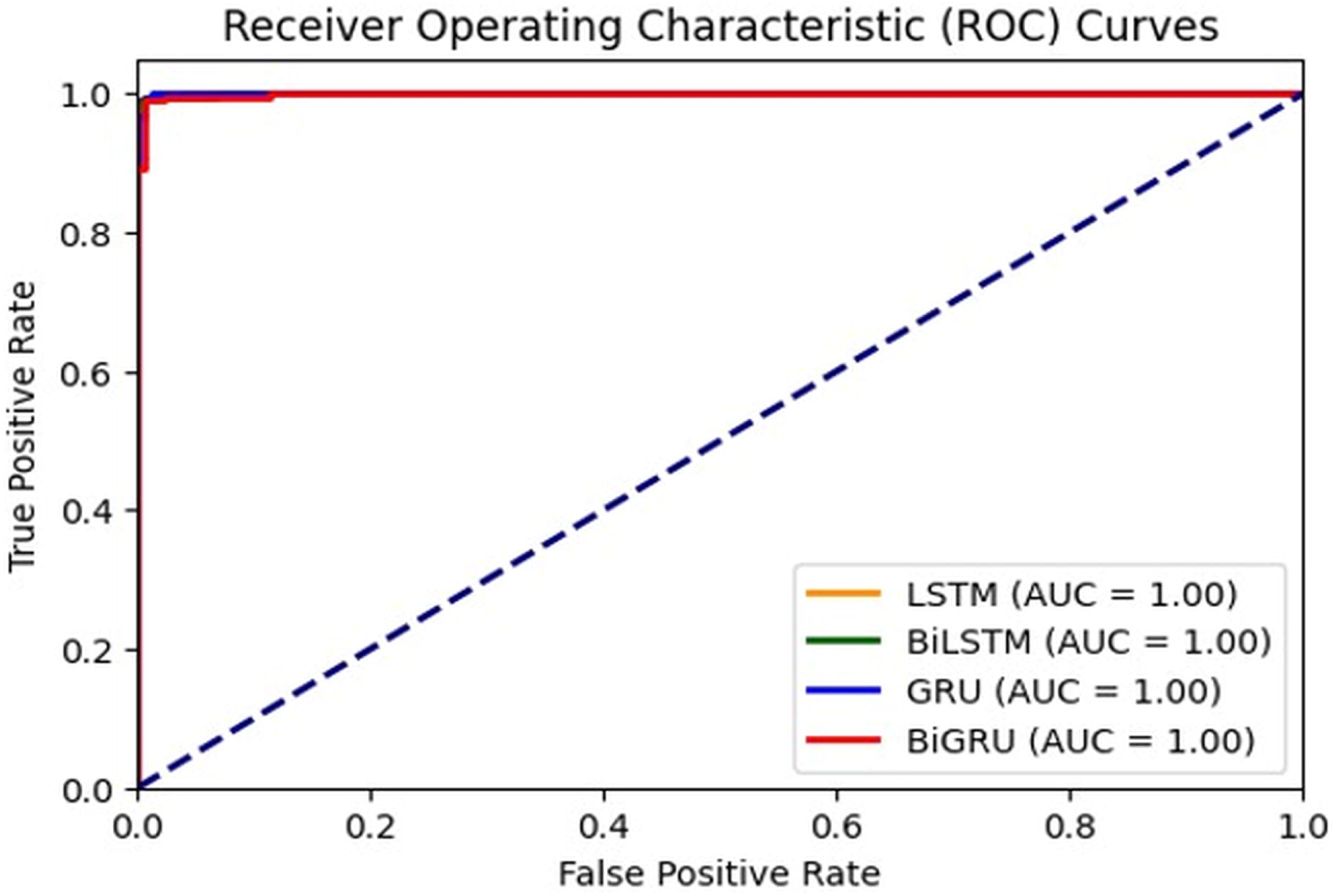

The unidirectional GRU model surpasses the LSTM, bi-LSTM, and its bidirectional variant, bi-GRU, in accuracy, recall, and F1 score. Although the other models yield comparable performance metrics, the GRU, being the more lightweight model in terms of parameter count and efficient training, effectively captures the pertinent sequential patterns in the data for females with superior performance and may be favoured over the alternative models.

## Discussion

The development of sex-stratified recurrent neural network designs for PD identification via VGRF data represents a significant advancement in deep learning techniques for diagnosing neurological illnesses. Our findings address critical deficiencies in the existing literature by demonstrating that sex-specific modeling, integrated with shorter time window analysis, yields highly effective diagnostic performance.

Our findings reveal notable differences in model performance between male and female groups, supporting the biological hypothesis that PD manifests differently across sexes. In male subjects, both unidirectional and bidirectional GRU models achieved comparable accuracy (98.5%), demonstrating outstanding recall (98.91%) and precision (98.64%) metrics. In contrast, for female subjects, the unidirectional GRU outperformed all other architectures, attaining an accuracy of 99.46%, a recall of 100%, and an F1 score of 99.48%. The models’ performance aligns with the findings of Haaxma et al.(20), who identified sex-based disparities in the onset and progression of PD. The sex-specific models likely revealed distinct biomechanical adaptations to dopaminergic degeneration that may have been masked in a combined model. This observation has important clinical implications, suggesting that diagnostic tools for PD should incorporate sex as a critical variable rather than treating it solely as a demographic factor.

The implementation of 5 s time windows represents a methodological advancement over previous studies that primarily utilized 10 s or longer time intervals. This reduces the time required for data collection and has practical implications for clinical implementation, as shorter assessment intervals are more feasible in time-constrained clinical settings and less burdensome for patients with mobility challenges. Despite the shorter duration, our models exhibited superior performance relative to those reported in the literature that employed longer windows, including the studies by Navita et al.(3) (97.5% accuracy), Mir et al.(7) (97.71% accuracy), and Setiawan et al.(13) (94.58% accuracy). This highlights the importance of using shorter time intervals in the investigation of alternative biomarkers of PD, as this approach may reveal hidden temporal dependencies that could be masked in longer time frames.

The comparative analysis of RNN variants revealed that GRU-based architectures consistently outperformed LSTM counterparts across both sex cohorts while requiring fewer parameters. For male subjects, the unidirectional and bidirectional GRU models achieved the same accuracy (98.5%), F1 score (98.78%), and other performance metrics. The unidirectional GRU significantly surpassed all other architectures, including its bidirectional counterpart, for female subjects. This performance pattern challenges the prevalent belief that bidirectional architectures inherently produce better outcomes for time-series classification tasks. The added complexity and computational expense of bidirectional processing did not yield significant performance enhancements, especially for female subjects. This finding suggests that forward sequential patterns in VGRF data are sufficiently distinctive for PD classification, whereas the reverse temporal context captured by bidirectional models offers minimal diagnostic value. The improved efficiency of GRU models is particularly relevant for potential use in resource-constrained environments or mobile applications, where computational efficiency directly affects accessibility. The comparable effectiveness of unidirectional GRUs to more complex architectures suggests that simplified models may be adequate for efficient PD screening, potentially enabling wider adoption of these technologies in primary care settings.

Despite the positive results, several limitations require further investigation. The dataset used for model development included subjects with moderate Parkinson’s disease severity (Hoehn and Yahr scores of 2 to 3), potentially limiting the models’ applicability to earlier disease stages where subtle gait abnormalities may be less apparent. Subsequent research should validate these methodologies on populations with earlier stages of Parkinson’s Disease to evaluate their efficacy for preclinical detection. Secondly, although sex stratification enhanced model efficacy, other demographic variables, including age-specific cohorts, cultural differences in gait patterns, and comorbidities influencing mobility, were not explicitly incorporated into the model. Investigating these supplementary stratification dimensions may enhance the personalization of PD detection algorithms. Ultimately, the existing models that concentrate solely on VGRF data, when integrated with additional biomarkers of PD, such as features derived from accelerometers, voice analysis, or digital biomarkers from smart devices, could establish more comprehensive detection systems. Future research should explore multimodal approaches that combine these complementary data sources to improve diagnostic accuracy.

## Conclusion

This study introduces a new methodology employing recurrent deep neural networks to create sex-specific models for identifying PD, laying the groundwork for the investigation of sex-specific diagnostic approaches. These models effectively identify temporal dependencies in VGRF signal data and exhibit notable classification performance when analyzed over short 5 s intervals. The rigorous data preprocessing ensures the exclusion of individuals under 50 years of age from the learning process, facilitating a more precise representation of the signal data from the older population. The implemented approach emphasizes the architectural benchmarks to consider when training recurrent neural networks for the utilization of VGRF signals in the diagnosis of PD. The model demonstrates that stratification based on demographic factors of sex and age can enhance the predictive accuracy for Parkinson’s disease. This methodology is applicable to other neurodegenerative disorders, such as Alzheimer’s disease, Huntington’s disease, and various gait-related conditions. The integration of supplementary biomarkers of Parkinson’s disease using this method can significantly improve the diagnostic process. This method simplifies the data transformation processes used in many studies, making it a viable choice for real-time clinical detection.

## Abbreviations

PD: Parkinson’s disease
VGRF: Vertical Ground Reaction Force
LSTM: Long Short Term Memory
GRU: Gated Recurrent Unit
bi-LSTM: bidirectional Long Short Term Memory
bi-GRU: bidirectional Gated Recurrent Unit
ML: Machine Learning
RNN: Recurrent Neural Networks
DL: Deep Learning
SVM: Support Vector Machines
k-NN: k-Nearest Neighbors
RFT: Random Forest Tree
ER: Ensemble Regressor
SMOTE: Synthetic Minority Oversampling Technique
NB: Naïve Bayes
GMM: Gaussian Mixture Models
LOOCV: Leave One Out Cross Validation
HC: Healthy Control
CWT: Continuous Wavelet Transformation
PCA: Principal Component Analysis
ROC: Receiver Operating Characteristic Curve
AUC: Area Under Curve.

## CRediT authorship contribution statement

Gurkirtan Singh:Writing – original

draft, Visualization, Methodology, Data curation, Conceptualization, Resources,

Formal Analysis, Anilendu Pramanik– Supervision, Data Curation, Validation, Resources, Conceptualization, Writing: review and editing.

## Data Availability

This study utilised publicly available dataset from the Gait in Parkinson’s Disease Database, accessible at PhysioNet (https://physionet.org/content/gaitpdb/1.0.0/). The dataset is freely available for download and use under PhysioNet’s Open Database License. No proprietary or restricted data were used in this analysis. All preprocessing scripts and model implementation code developed for this study are available from the corresponding author upon reasonable request.

## Acknowledgements

I would like to sincerely thank Dr. Anilendu Pramanik for his unwavering support and guidance throughout this research. His expertise in biomechanics, research methodology, research paper writing, and the publication process greatly enhanced the quality and rigor of this work. I am deeply grateful for his readiness to provide insightful advice whenever I encountered challenges.

I also wish to express my gratitude to Guru Nanak Dev University and Dr. for providing the academic environment and resources that enabled me to undertake this research. It was through the opportunities and support of the university that I was able to connect with Dr. Pramanik and benefit from his mentorship.

